# Systematic review of pro-equity strategies to improve vaccination among priority populations

**DOI:** 10.1101/2025.06.03.25328765

**Authors:** Adeline Tinessia, Majdi M. Sabahelzain, Catherine King, Saman Khalatbari-Soltani, Praveena Gunaratnam, Ibrahim Dadari, John Carlo Lorenzo, Soumyadeep Bhaumik, Meru Sheel

**Affiliations:** Sydney School of Public Health, Faculty of Medicine and Health, The University of Sydney, New South Wales, Australia; Sydney Infectious Diseases Institute, Faculty of Medicine and Health, The University of Sydney, New South Wales, Australia; Clinton Health Access Initiative (CHAI), Vientiane Capital, Lao People’s Democratic Republic; Coverage & Equity Unit, Immunization Section, PG-Health, UNICEF Headquarters, New York, NY, USA; Meta-research and Evidence Synthesis Unit, The George Institute for Global Health, Australia; Faculty of Medicine and Health, University of New South Wales, Sydney, New South Wales, Australia

## Abstract

**Background/Objectives:** The importance of pro-equity strategies in addressing disadvantages that people and communities face due to their gender, migration status, ethnicity, disability, and place of residence is increasingly being recognised, but analysis of empirical evidence on how they improve vaccination in these priority groups is limited. This systematic review aims to fill this gap.

**Methods:** Standard evidence synthesis methods were employed, with searches conducted in four major bibliographic databases in March 2025. Studies were included if they reported on strategies aimed at improving vaccination uptake, coverage, and/or timeliness among priority groups disadvantaged by gender, migration status, disability, ethnicity, or geographical location, with no exclusions based on language, time, or type of vaccination. Excluded studies were those without explicit intervention or outcome data related to vaccination and those not focused on the identified priority groups or settings. A thematic analysis was conducted to map strategies for immunisation programmatic areas. Strategies were also mapped to the Tanahashi coverage framework, which identifies how pro-equity strategies addressed bottlenecks in the delivery of immunisation programs.

**Results:** From 20,812 records retrieved, we identified 59 studies showing pro-equity strategies that improved vaccination uptake or coverage (n=54) and timeliness (n=6). Twenty-five pro-equity vaccination strategies under five immunisation system domains were studied. Thirty-nine studies were conducted in high- and middle-income countries. Most (56%) focused on community-oriented strategies and improving acceptability (63%). Few targeted upstream system-level barriers, such as policy and governance (n=7), availability (n=5), or effectiveness (n=10) of immunisation service delivery. No single strategy or approach was universal across the priority groups. However, some approaches were more common in specific populations and settings.

**Conclusions:** This review synthesises strategies to address vaccination inequities among underserved populations. Achieving vaccination equity will require context-specific approaches to address individual and systemic barriers across community engagement, service delivery, information systems, and policy and governance.

**Funding:** None

## Introduction

The Immunization Agenda 2030 (IA2030) highlights the importance of “coverage and equity” as a strategic priority area to ensure all people are protected by vaccination, regardless of location, age, socioeconomic position or gender (1). Immunisation inequities, defined as systematic differences in vaccination that leave certain individuals or population groups at risk of being unvaccinated or under-vaccinated, can exist in many forms, including by age, geography and socioeconomic position (1).

The COVID-19 pandemic brought vaccine inequities to the global forefront (2), and they remain an issue. For example, the numbers and distribution of zero-dose children (defined as children who have not received a single dose of the Diphtheria-Tetanus-Pertussis-DTP vaccine) show deep vaccine inequities across countries and regions (3,4), with 59% of zero-dose children in 2023 living in just 10 countries: India, Nigeria, Ethiopia, Democratic Republic of Congo, Sudan, Indonesia, Yemen, Afghanistan, Angola, and Pakistan (4).

Social, economic, and health system factors shape immunisation inequities. Evidence suggests that vaccination rates are often lower in certain groups, including women and gender minorities; migrants and mobile populations; people living with disabilities; ethnic minorities; and rural or urban poor populations (3–6). Women and men have different experiences in access to vaccination, influenced by the social roles and norms that affect health decisions. For example, a 52-country study found that median DTP3 coverage was 12% greater among children of women with the highest social independence compared with children of women with the lowest (6). Migrants and mobile populations face healthcare discrimination, language barriers, or lack of trust in local health systems, with a scoping review finding that 70% of studies noted lower rates of immunisation among migrants (7). People living with disabilities often face physical, social, and systemic barriers to healthcare, which can limit access to vaccination (8,9) and increase their vulnerability to diseases. A study of 97 low- and middle-income countries (LMICs), significant differences in vaccination coverage existed between urban and rural areas, where the pooled prevalence of zero-dose children was 4.7% for rural areas, 12.6% for urban poor and 6.5% among urban non-poor (10). A systematic review of 64 countries found that the median gap between the highest and lowest zero-dose prevalence ethnic groups in all countries was 10% (11). These barriers often overlap across individuals, populations, and contexts.

Acknowledging these known gaps, there is a need to understand, design, and implement effective strategies to reduce immunisation inequities (12). Pro-equity vaccination strategies are targeted approaches to improve vaccination among underserved and priority groups, by increasing utilisation and adopting a health system strengthening approach (13–15). Recent publications identified pro-equity vaccine strategies in some Gavi-supported countries (13–15). However, there is limited evidence on the effectiveness of such strategies in reducing coverage gaps. This systematic review aims to identify pro-equity strategies used to improve vaccination amongst priority groups, i.e. women and gender minorities, migrants and mobile population, people living with disabilities, ethnic minorities and urban and rural population, and to inform how to address global gaps in vaccination coverage and equity. Specifically, we examined the impact of these strategies on vaccination uptake, coverage, and/or timeliness.

## Methods

This review followed the Preferred Reporting Items for Systematic Reviews and Meta-Analyses (PRISMA) 2020 checklist (Appendix A) (16). The protocol was registered on the International Prospective Registration of Systematic Reviews (PROSPERO 2023 CRD42023449067) (http://www.crd.york.ac.uk/PROSPERO) (17).

### Eligibility criteria

Studies were included if they reported on pro-equity strategies targeting priority groups and vaccine-related outcomes, including change in vaccine uptake, coverage, and/or timeliness. Priority groups included sub-populations disadvantaged due to their gender, migration status, disability, ethnicity, or geographical location (urban/rural). Gender was defined as socially constructed roles, norms and behaviours that societies consider appropriate, which shape relationships within and between gender groups (1). Migration status included refugees, asylum seekers, and transient populations (18).

Disability was defined as an umbrella term for impairments, activity limitations, and participation restrictions, reflecting the negative interaction between an individual’s health condition and contextual factors (19). Geographically disadvantaged included urban populations living in slums, informal or high-density areas, and rural populations with limited health care access. We defined pro-equity strategies as those that promoted immunisation access and uptake among priority groups (13).

Randomised controlled trials, quasi-experimental studies, observational studies, cross-sectional studies, pre-post intervention studies, controlled community trials, cluster-randomised trials, mixed methods studies and qualitative studies were included. No language-based or time-based exclusions were made. All types of vaccinations are included. Studies were excluded if they were animal studies, ecological studies, case series, commentaries, reviews, letters or news articles; if they did not explicitly discuss interventions aimed at increasing vaccination uptake, coverage or timeliness; if they did not explicitly have outcome data on vaccination uptake, coverage, or timeliness; or if they did not address one of the priority groups or settings of interest.

### Search strategy

Four databases were searched: Ovid MEDLINE (1946 to March 11, 2025), Ovid Embase Classic (1947 to 1973) + Ovid Embase (1974 to March 19, 2025), OVID Global Health (1910 to Week 11 2025) and Scopus (1823 to March 2025). The detailed strategy is available in Appendix B. An updated search was conducted on 20 March 2025. Grey literature was searched using Google Scholar. References of included studies were also snowballed for additional articles.

### Study selection

Search results were first deduplicated using EndNote and manually checked by the first author (AT). The titles and abstracts of all studies were screened by two reviewers (AT screened all records, JCL and MMS conducted secondary screening of all records, with screening workload shared between them). Full texts were screened independently by AT, JCL and MMS. Disagreements were resolved through consensus with the authorship team. Rayyan was used to manage the review process and record decisions.

### Data extraction

Two reviewers (AT, CL, MMS, SKS, PG, ID) independently extracted data from all included studies. Results were compared by AT, and disagreements were discussed until a consensus was reached. Data were extracted across five domains: study identifiers, study specifics, information on participants, immunisation pro-equity strategy information, and limitations. Microsoft Excel was used for data extraction.

### Risk of Bias Assessment

The Mixed Methods Appraisal Tool (MMAT) version 2018 was used to assess and evaluate the trustworthiness, relevance, and results D of the included studies (20). MMAT evaluated studies into high (scores of 4-5/5), moderate (scores of 3/5), and low (scores of 1-2/5) quality. Two researchers assessed each study independently (AT, CL, MMS, SKS, PG, ID), and discrepancies between reviewers were resolved through discussion.

### Data analysis

Priority group characteristics, including country income level, outbreak setting, routine immunisation status, use of multiple interventions in the same study and vaccine outcomes, were tabulated into frequency data. For ease of presentation, data on uptake and coverage were combined.

We used thematic analysis to identify and categorise pro-equity immunisation interventions. Intervention descriptions were inductively coded by two reviewers (AT, MMS). Unique codes were classified into discrete strategies, based on similarities in intervention design, delivery, and intent (Supplementary Table 1) and synthesised into five higher-order immunisation system domains: community-oriented, service delivery, health worker capacity oriented, immunisation or health management information system, and policy and governance (Supplementary Table 2).

We deductively classified the strategies using the Tanahashi framework (21), focusing on five domains of health service coverage: availability, accessibility, acceptability, contact, and effectiveness (Supplementary Table 3). Using the Tanahashi approach allowed for the identification of how pro-equity strategies addressed bottlenecks in the delivery of immunisation programs.

## Results

Of the 20,812 identified records, 10,037 records underwent title and abstract screening, 231 underwent full-text review, resulting in 59 studies which met the eligibility criteria (Figure 1).

**Figure 1.**
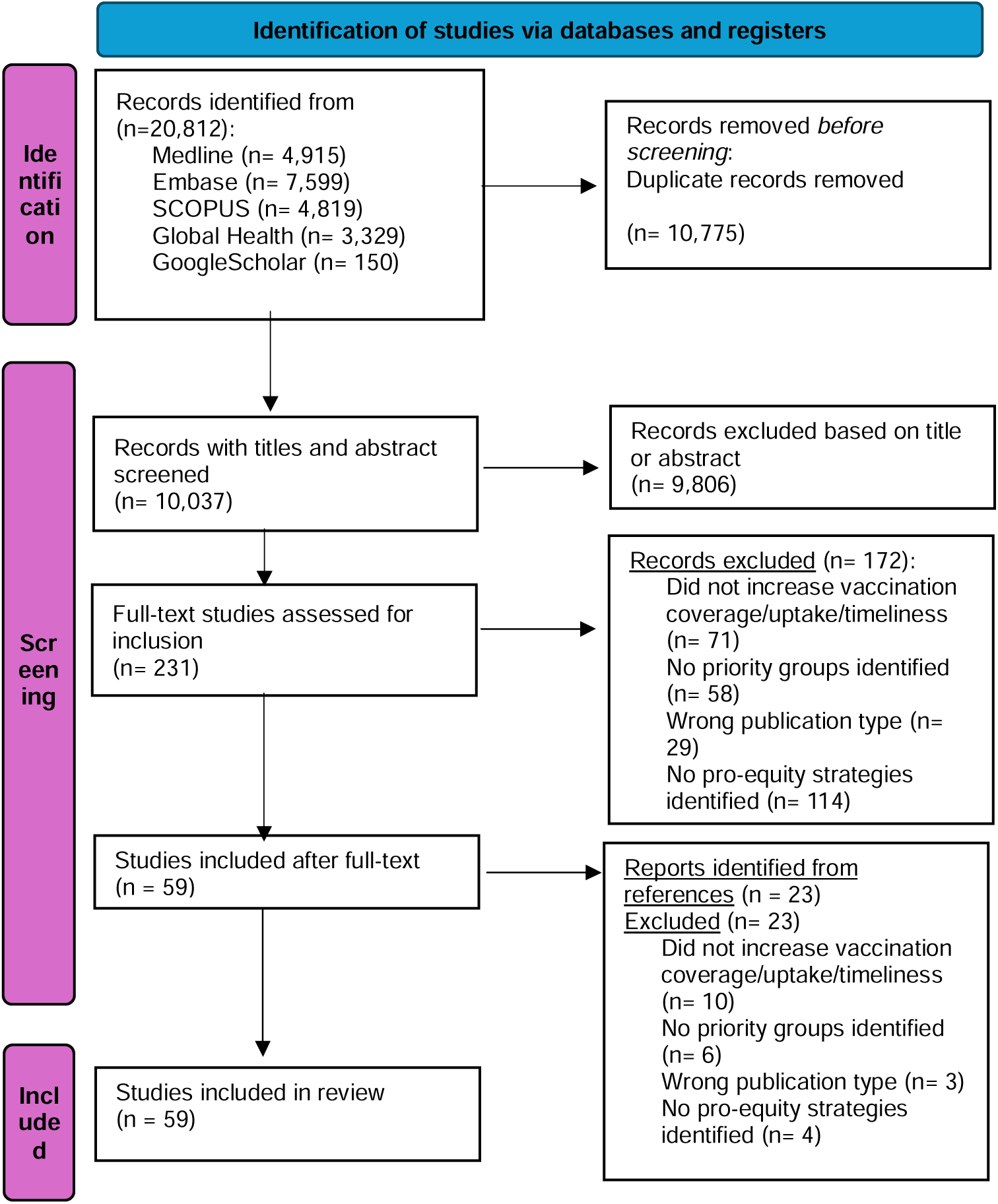
PRISMA flow diagram for the identification, screening and inclusion procedure of eligible articles found in the literature

### Characteristics of the included studies

Of the 59 included studies, 39 (66%) were conducted in high-income countries (HICs) and upper middle-income countries (UMICs), and the remaining 20 (34%) in LMICs or low-income countries (LICs). Over half (32/59; 54%) targeted children under 18 years of age. Thirty-one (53%) studies focused on improving vaccination outcomes for routine immunisation (Table 1). Three (5%) studies focused on improving outcomes for COVID-19 vaccinations, 3 (5%) for influenza vaccination, and 14 (24%) for HPV vaccination. Twenty-two (37%) were pre-post studies, 16 (27%) observational studies, 11 (19%) randomised controlled trials (RCTs) including group/cluster RCTs, 6 (10%) mixed methods, 3 (5%) case studies, and one a non-randomised controlled trial (Table 2). Twenty-four (41%) were from the United States of America (USA), 4 (7%) were from India and Nigeria each, 3 (5%) from Pakistan, two from Australia, Kenya, United Kingdom (UK), and Hong Kong each, and one study each from Namibia, South Africa, Nepal, France, Switzerland, Denmark, Spain, Rwanda, Thailand, Somalia, Brazil, Uganda, South Sudan, Colombia, and Japan (Figure 2).

**Figure 2.**
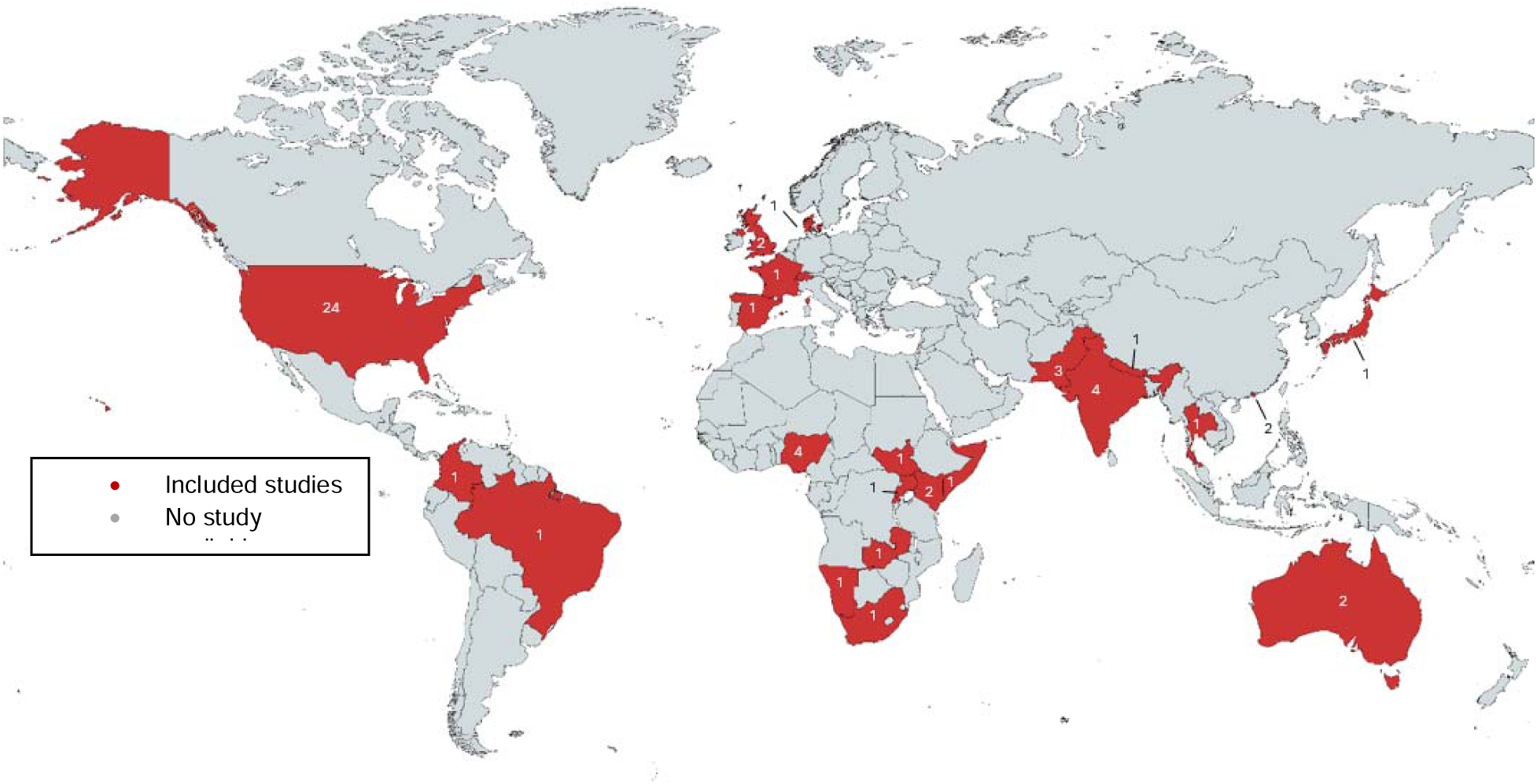
Map depicts the distribution of included studies. Numbers depict the number of studies per country. The boundaries and designation shown on this map are software-generated and do not imply any positioning or expression of opinion by the authors or their institutions regarding the legal status of any country, territory, city, or area, or the delimitation of its frontiers and boundaries.

**Table 1.**
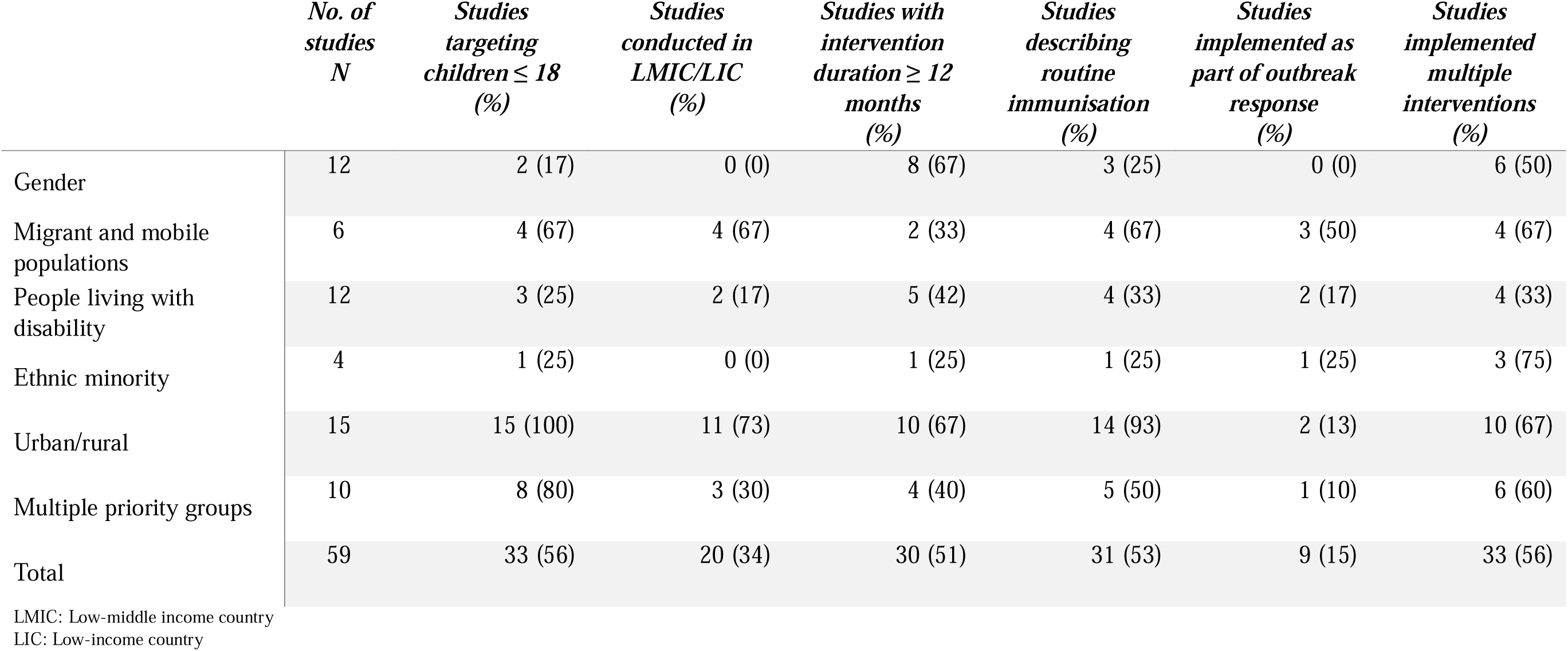
Summary description of studies describing pro-equity strategies to increase vaccine coverage, uptake, or timeliness.

|  | *No. of*<br>*studies*<br>*N* | *Studies*<br>*targeting*<br>*children ≤ 18*<br>*(%)* | *Studies*<br>*conducted in*<br>*LMIC/LIC*<br>*(%)* | *Studies with*<br>*intervention*<br>*duration ≥ 12*<br>*months*<br>*(%)* | *Studies*<br>*describing*<br>*routine*<br>*immunisation*<br>*(%)* | *Studies*<br>*implemented as*<br>*part of outbreak*<br>*response*<br>*(%)* | *Studies*<br>*implemented*<br>*multiple*<br>*interventions*<br>*(%)* |
| --- | --- | --- | --- | --- | --- | --- | --- |
| Gender | 12 | 2 (17) | 0 (0) | 8 (67) | 3 (25) | 0 (0) | 6 (50) |
| Migrant and mobile<br>populations | 6 | 4 (67) | 4 (67) | 2 (33) | 4 (67) | 3 (50) | 4 (67) |
| People living with<br>disability | 12 | 3 (25) | 2 (17) | 5 (42) | 4 (33) | 2 (17) | 4 (33) |
| Ethnic minority | 4 | 1 (25) | 0 (0) | 1 (25) | 1 (25) | 1 (25) | 3 (75) |
| Urban/rural | 15 | 15 (100) | 11 (73) | 10 (67) | 14 (93) | 2 (13) | 10 (67) |
| Multiple priority groups | 10 | 8 (80) | 3 (30) | 4 (40) | 5 (50) | 1 (10) | 6 (60) |
| Total | 59 | 33 (56) | 20 (34) | 30 (51) | 31 (53) | 9 (15) | 33 (56) |

LMIC: Low-middle income country LIC: Low-income country

**Table 2.**
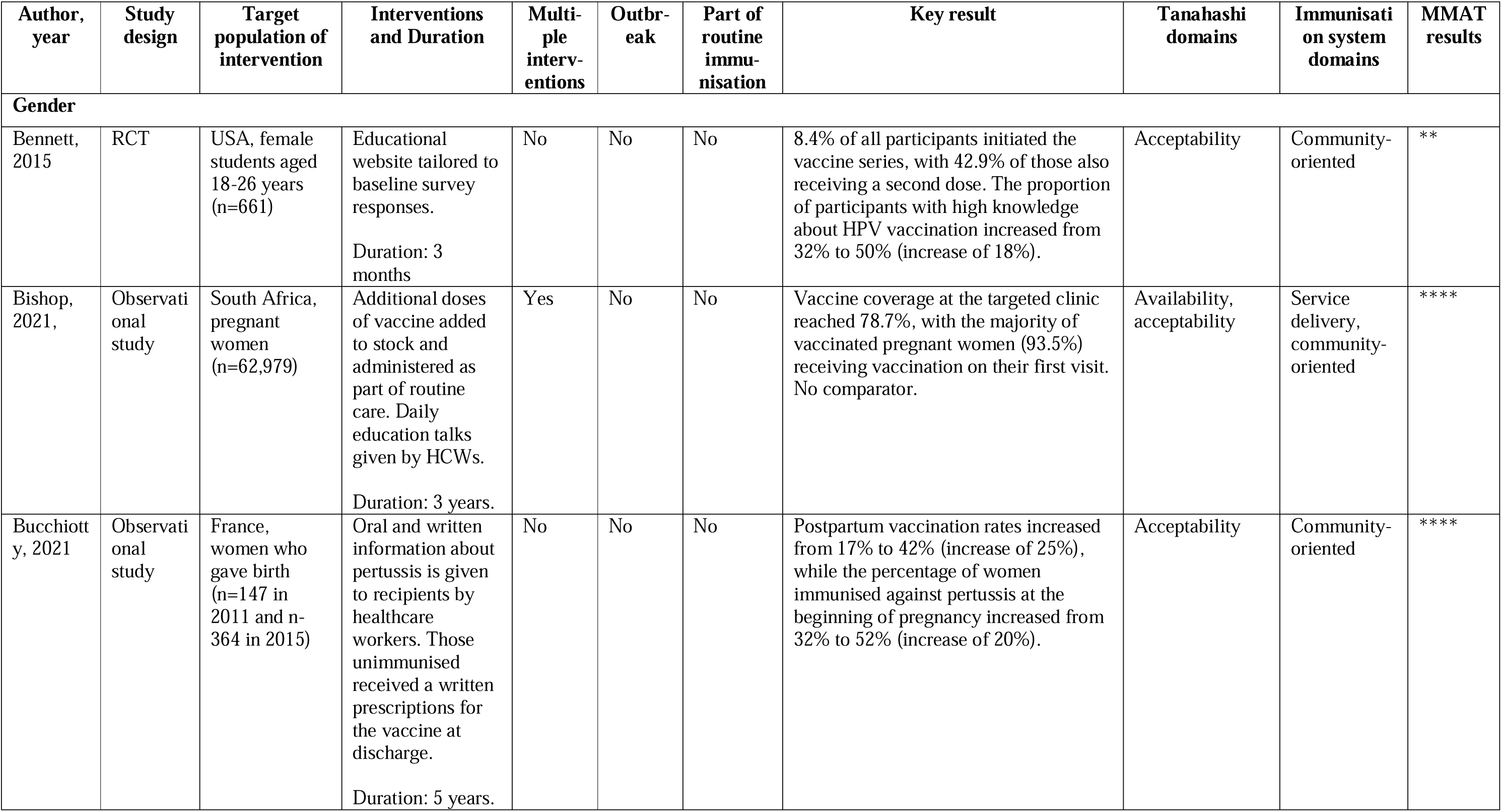
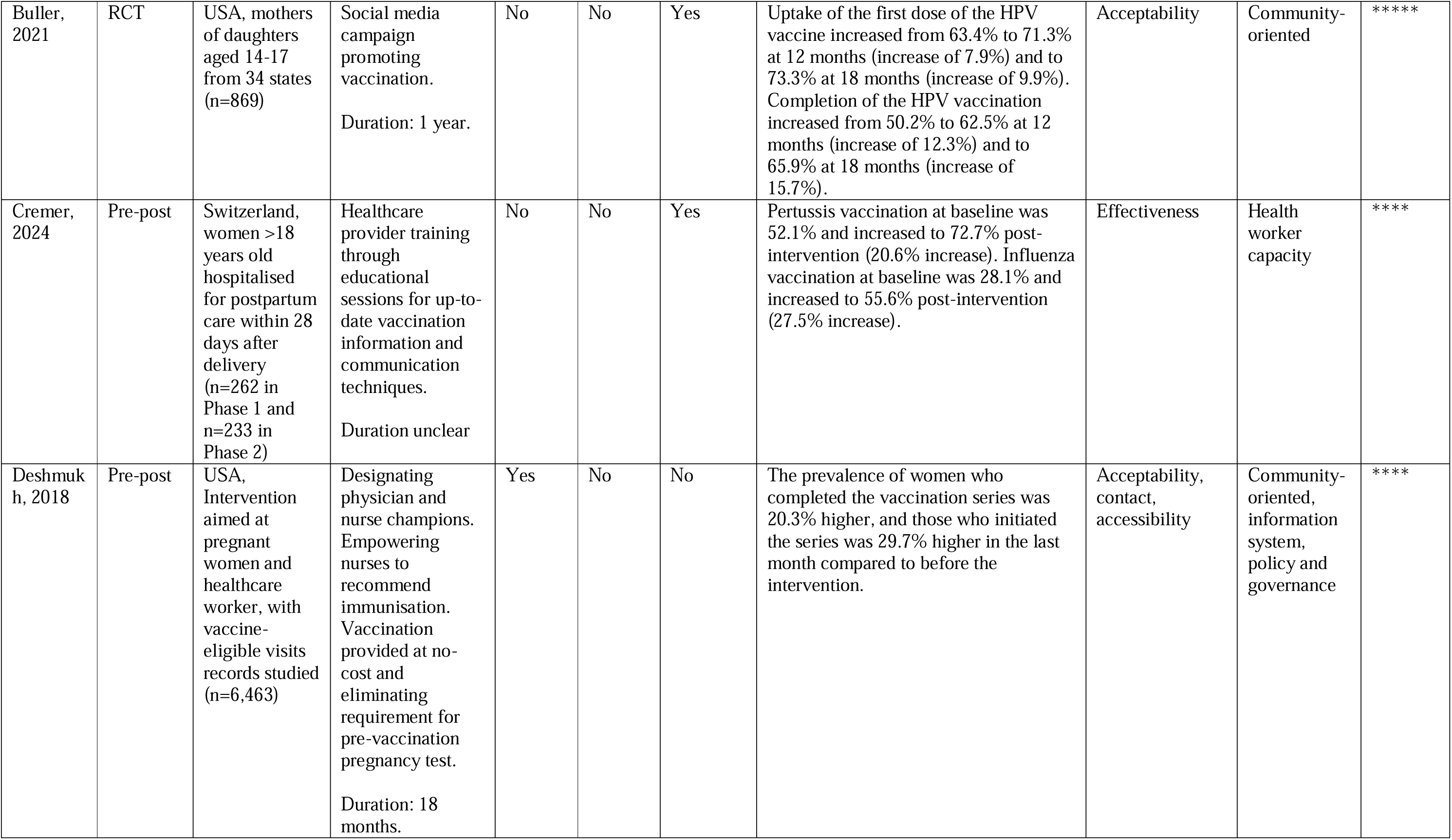
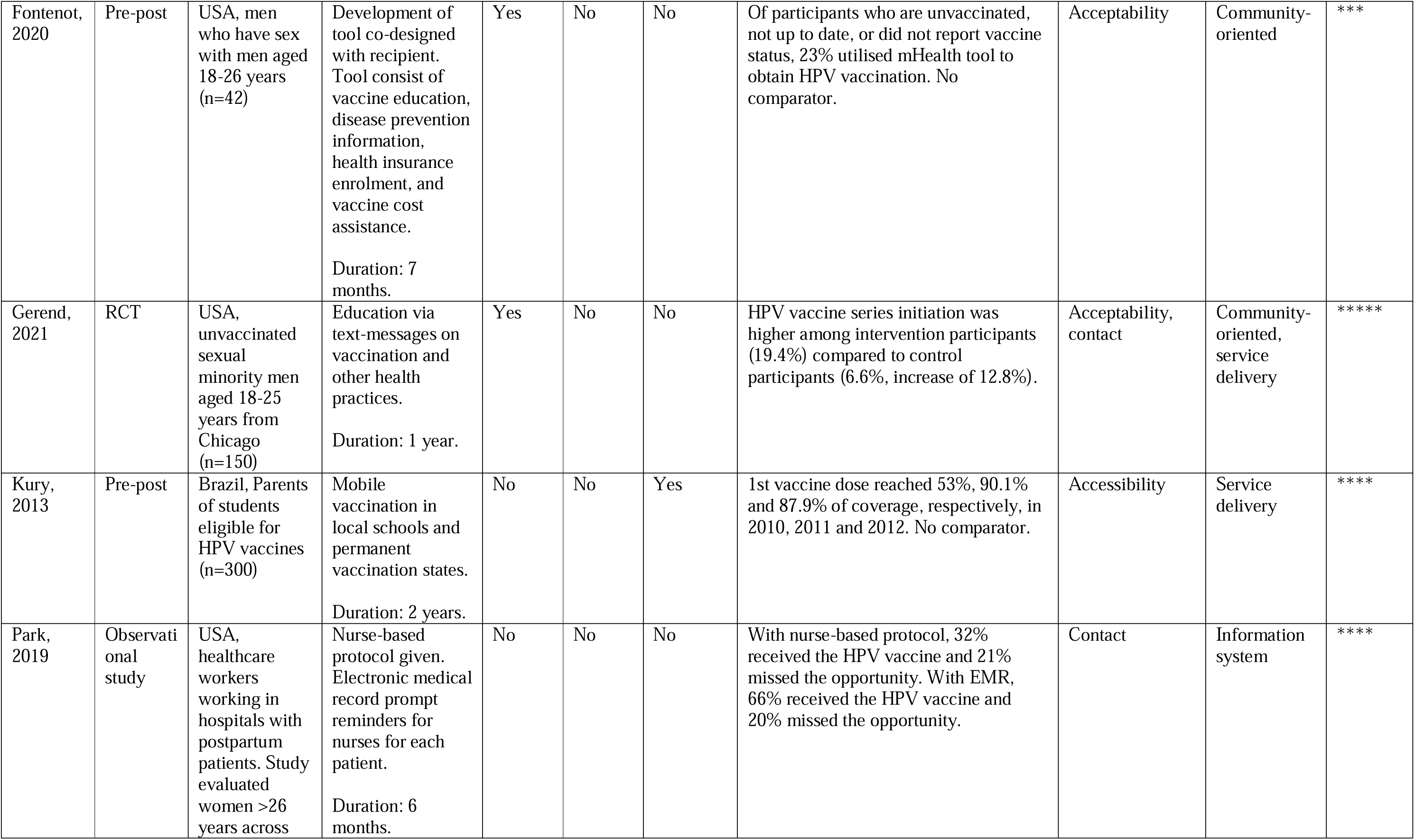
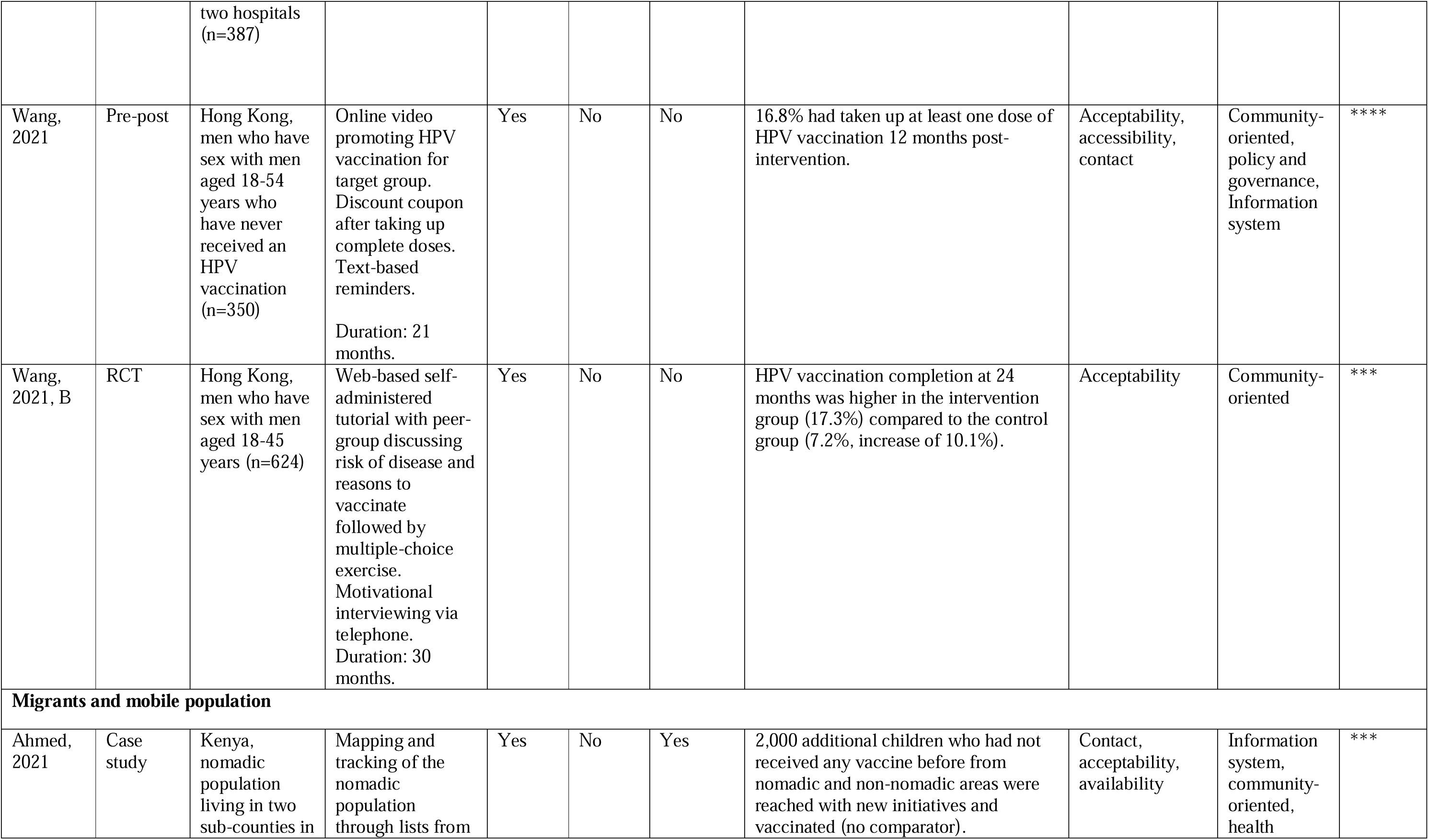
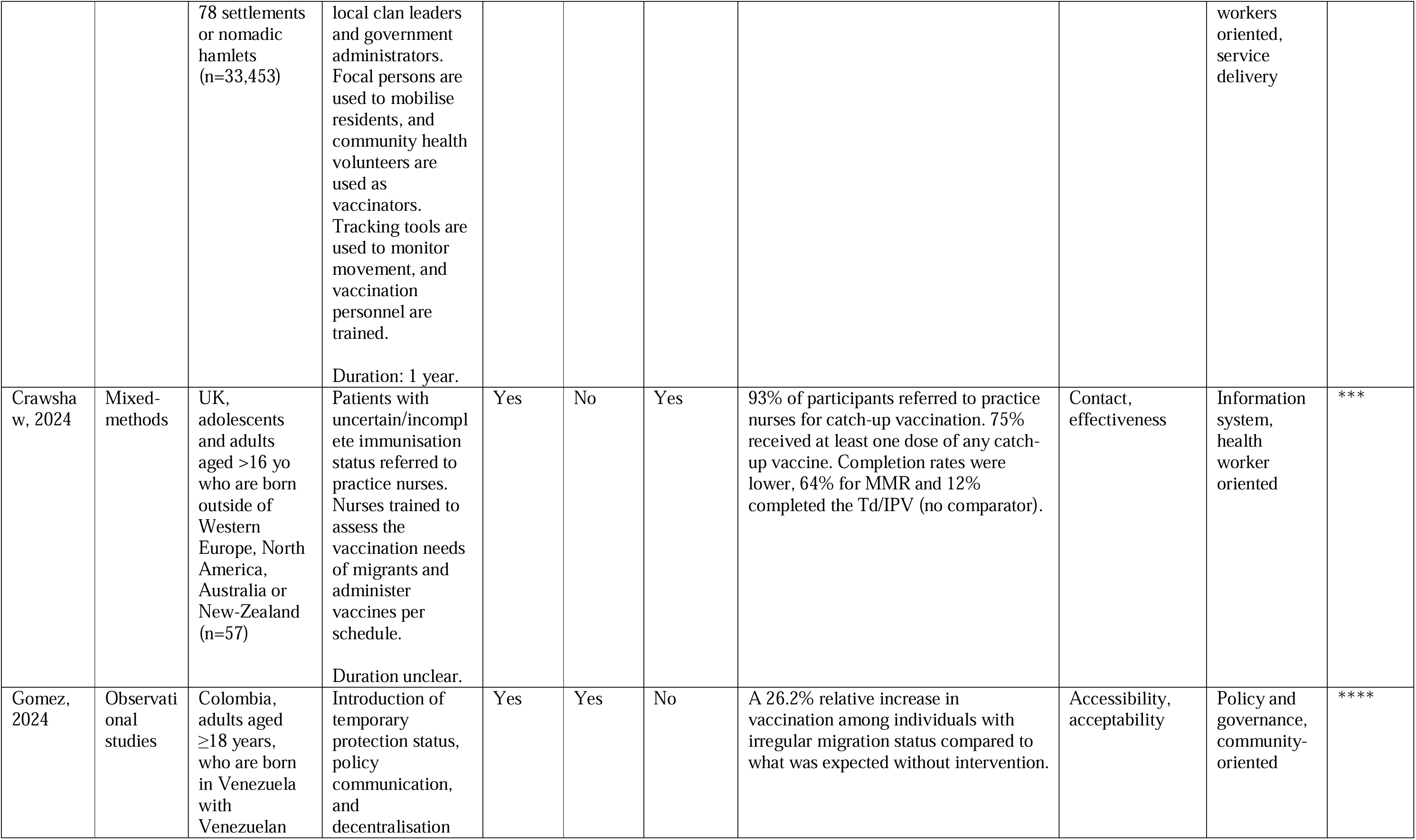
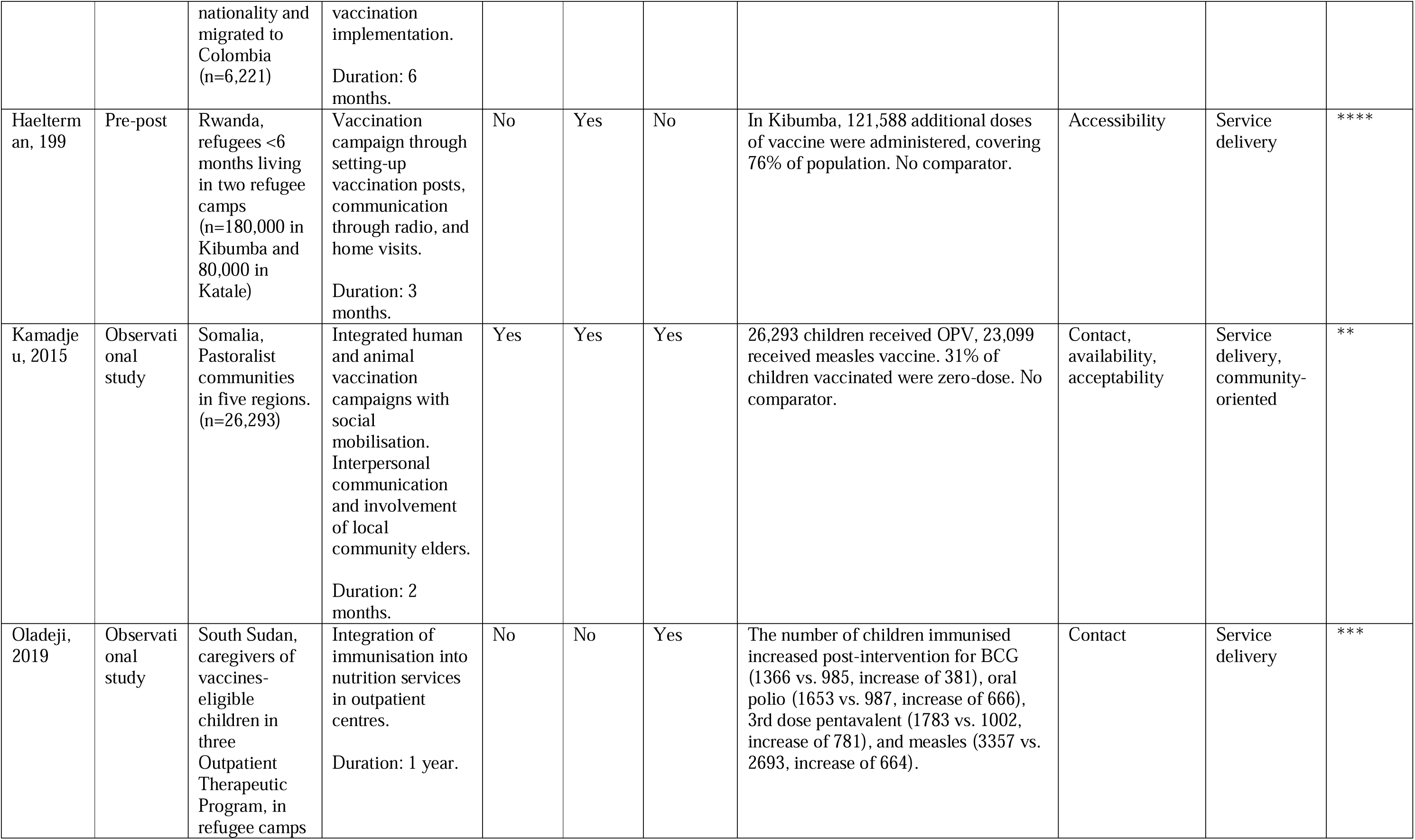
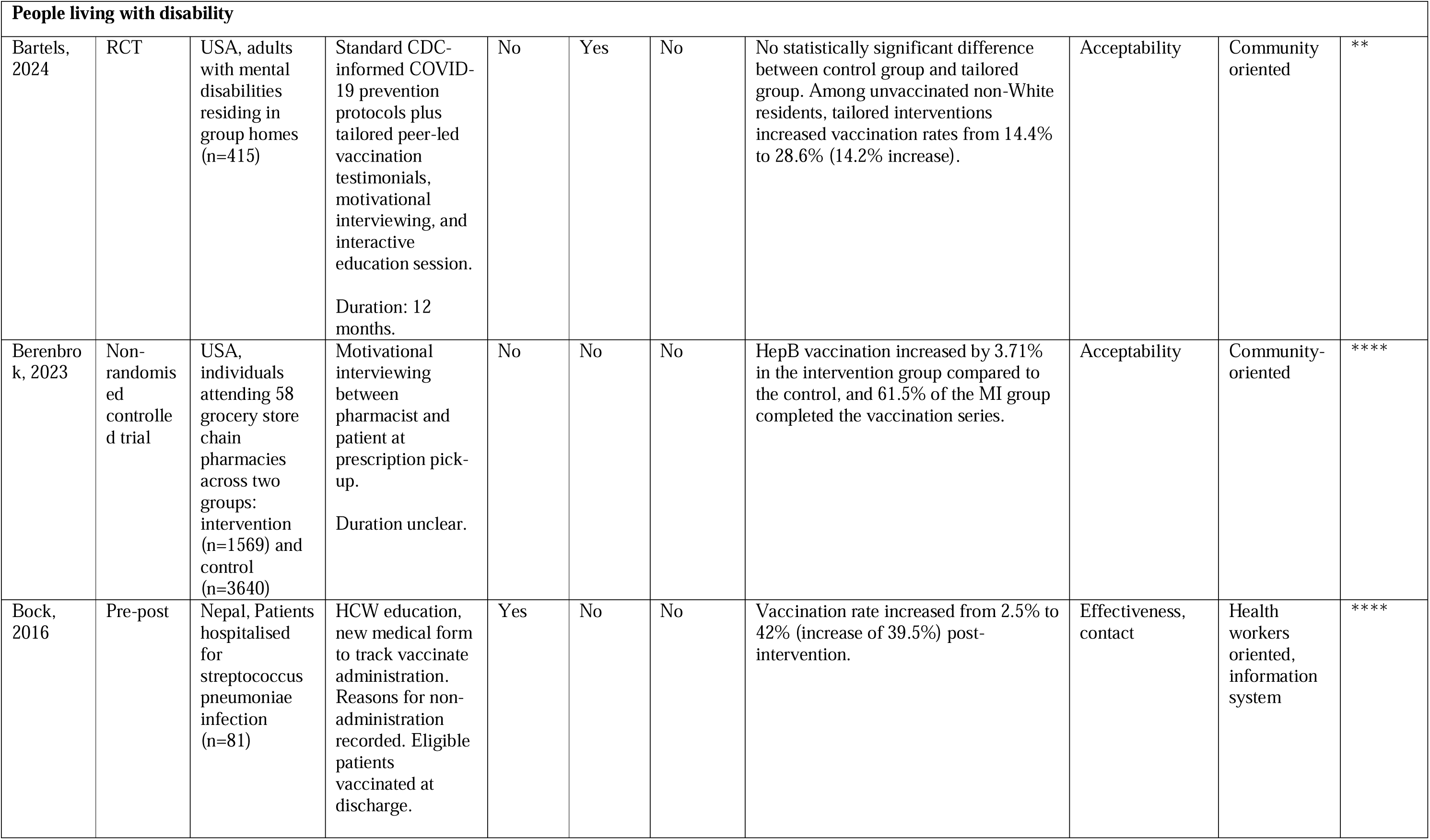
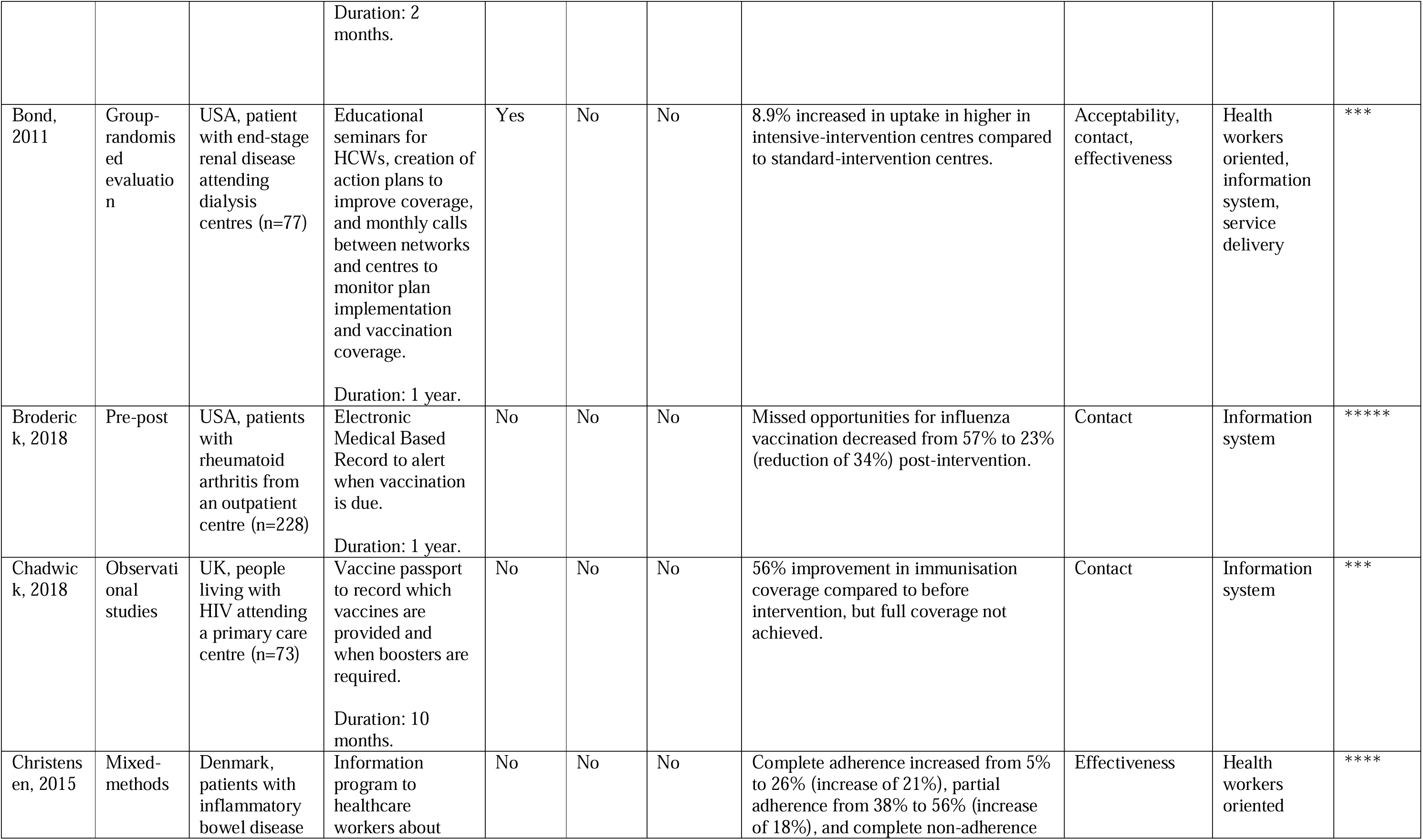
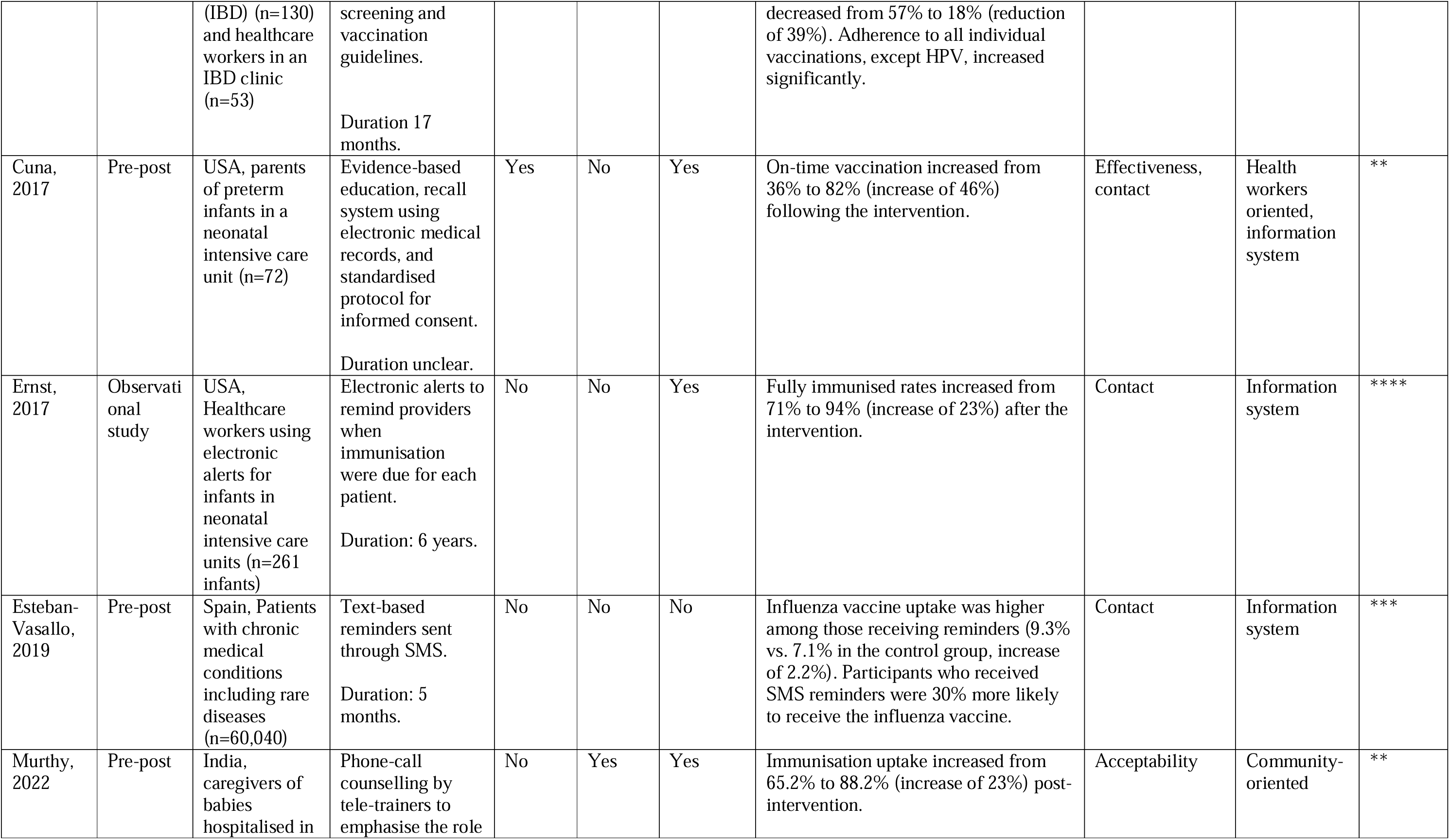
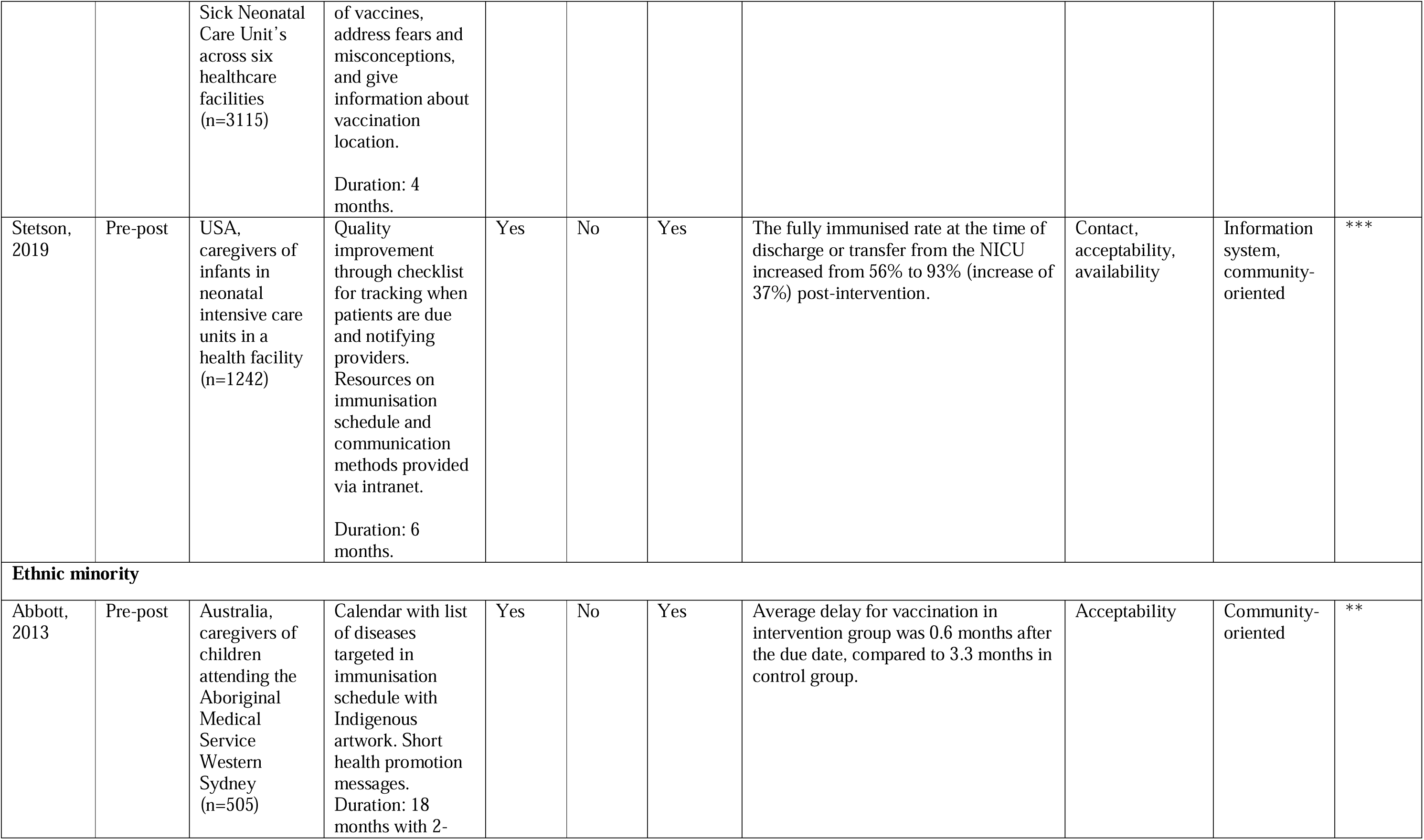
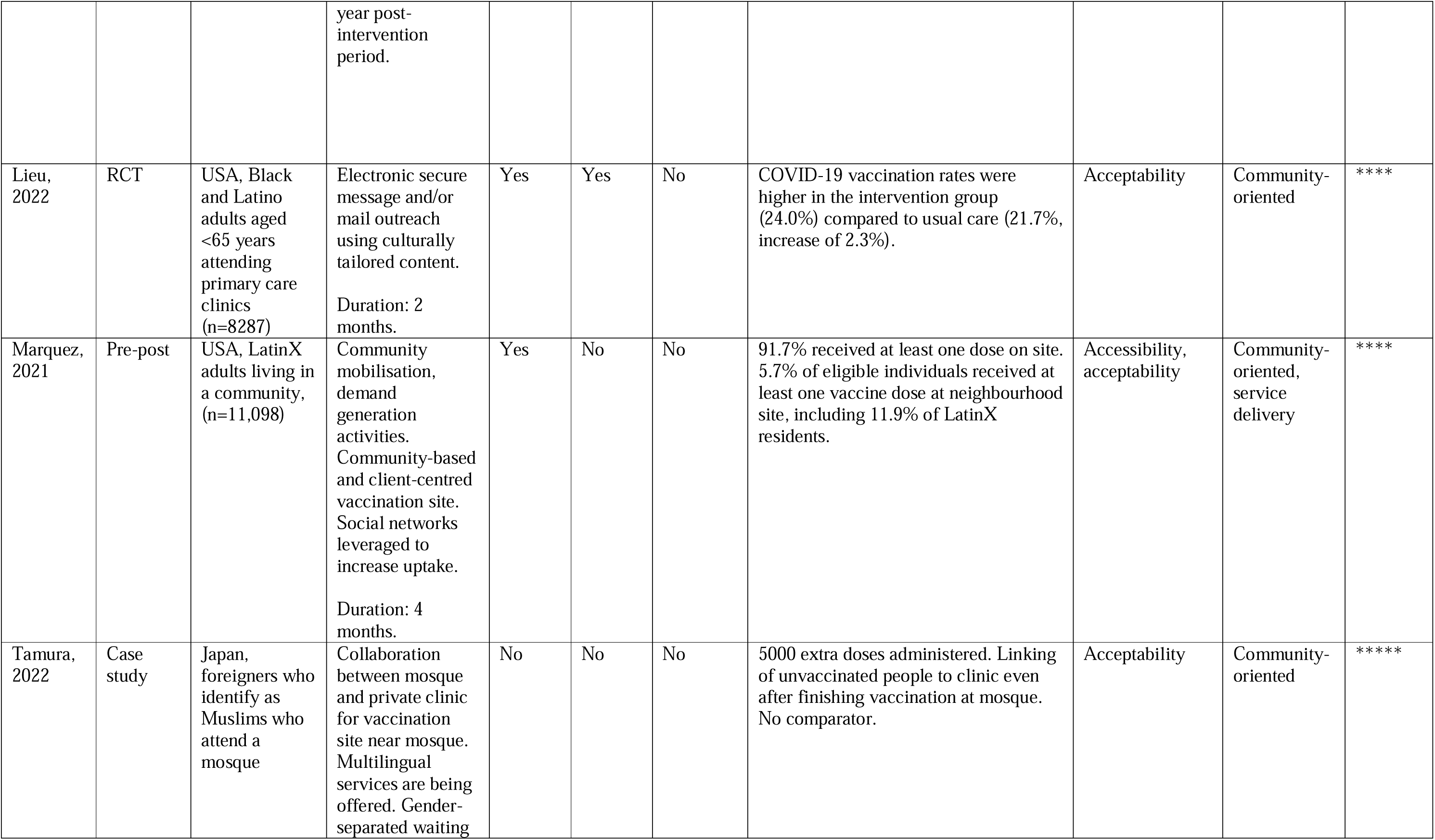
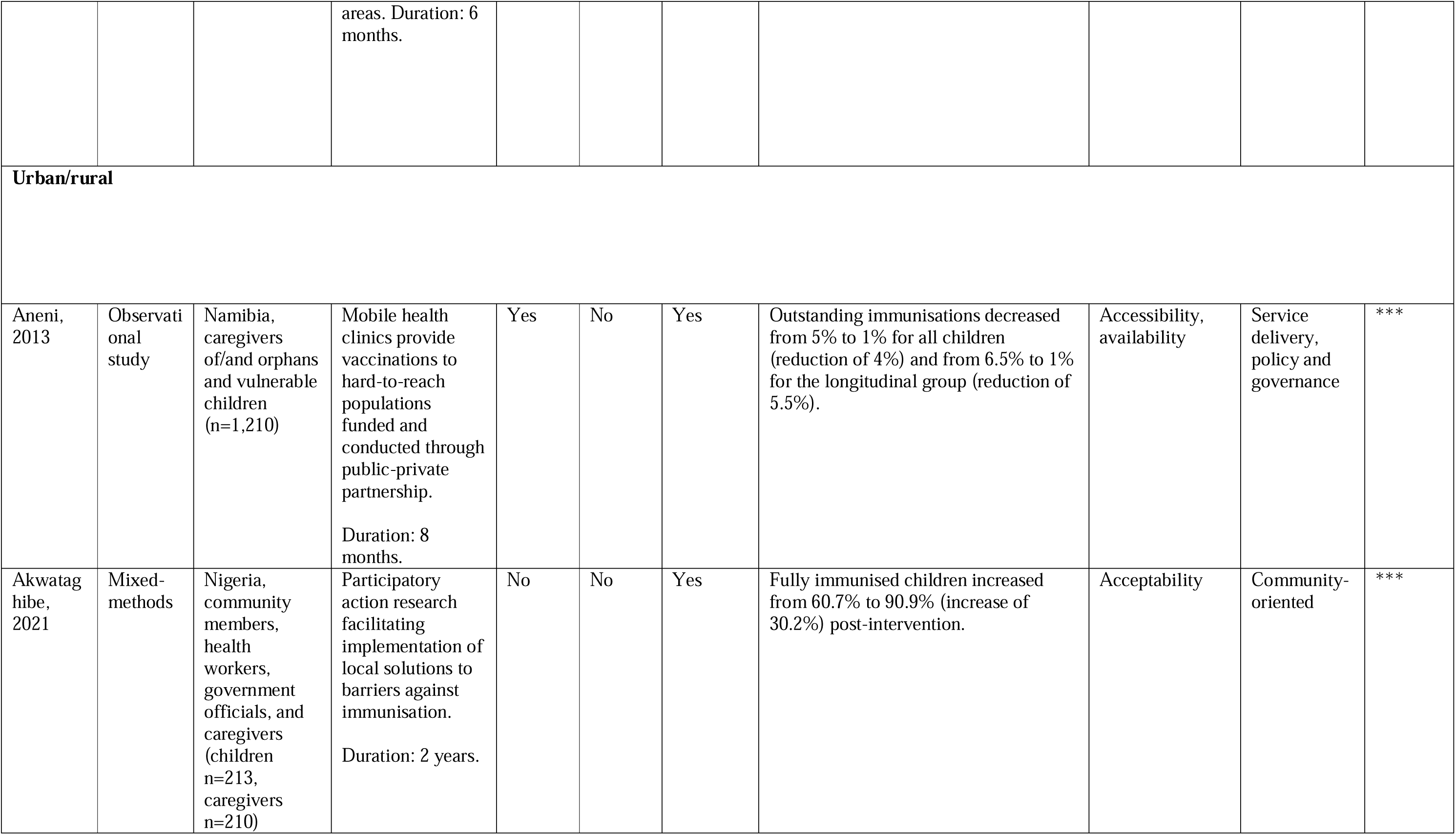
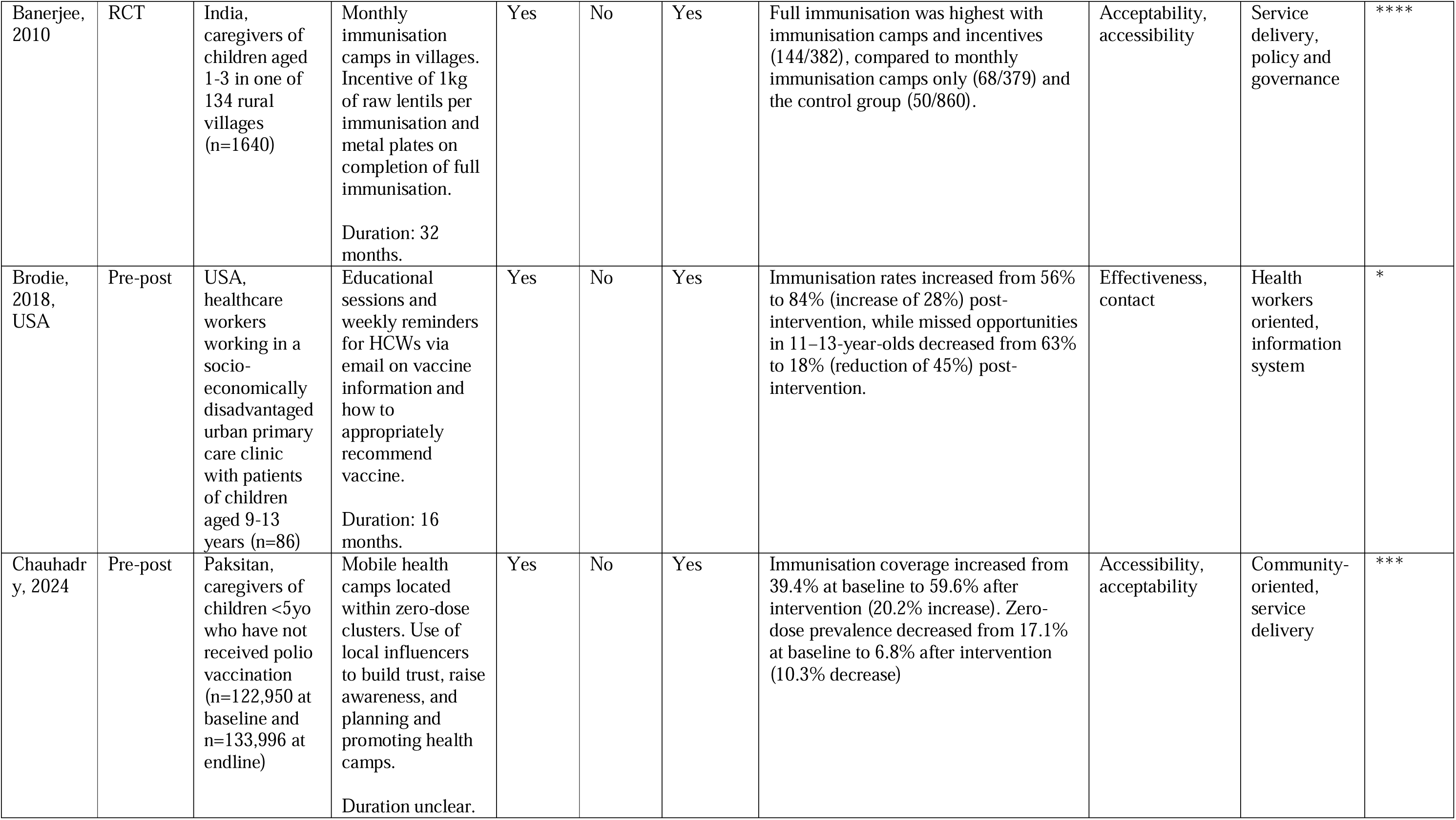
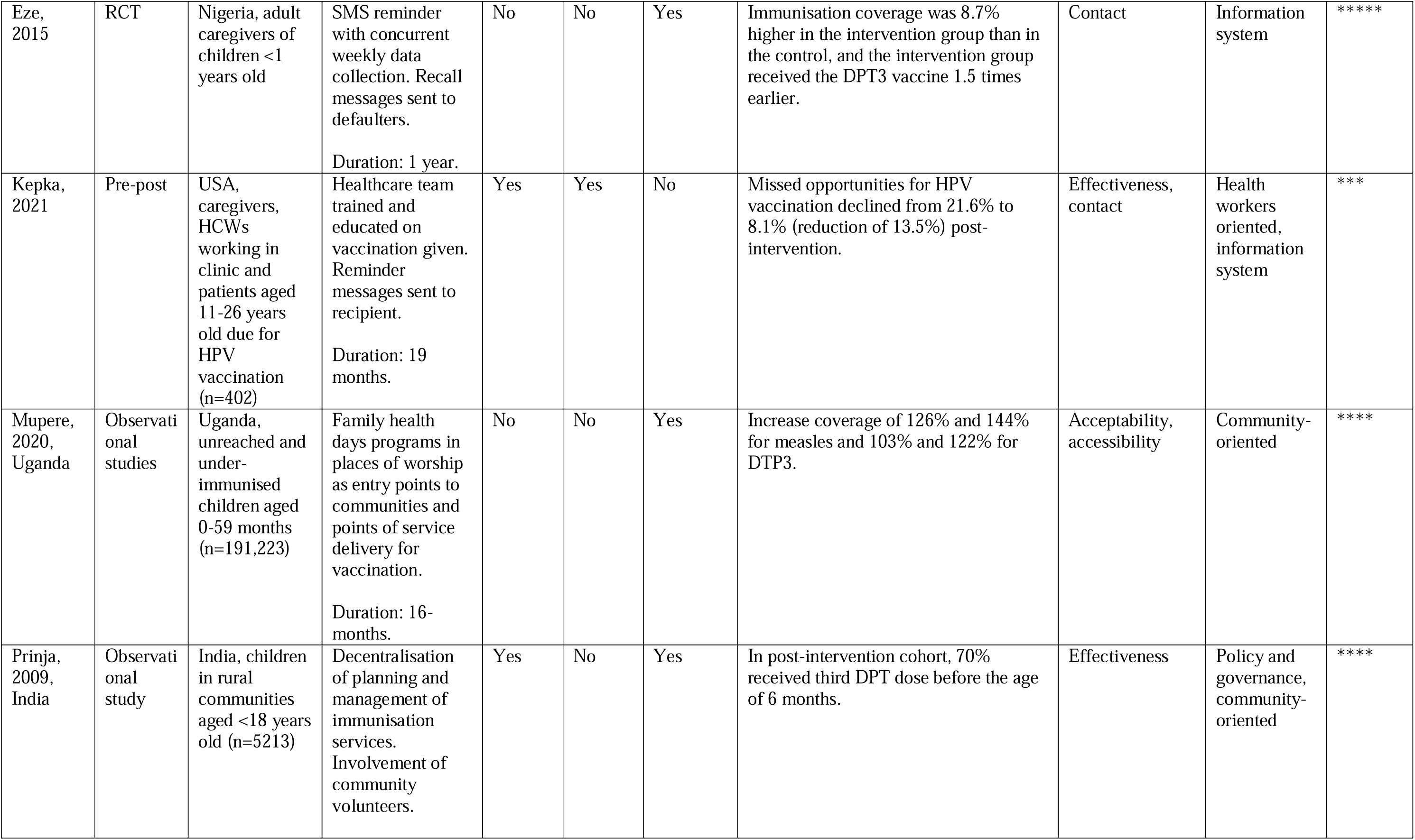
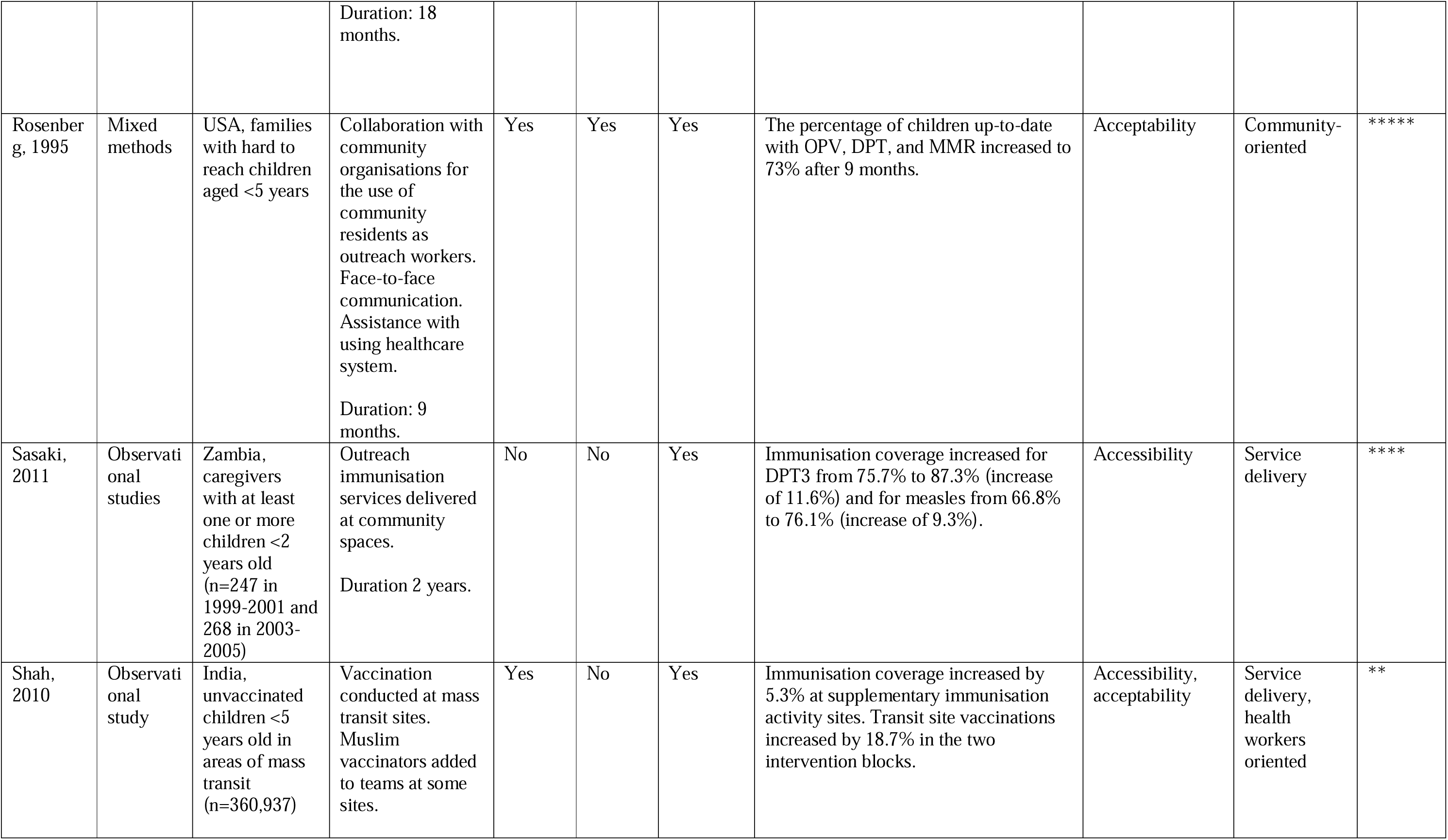
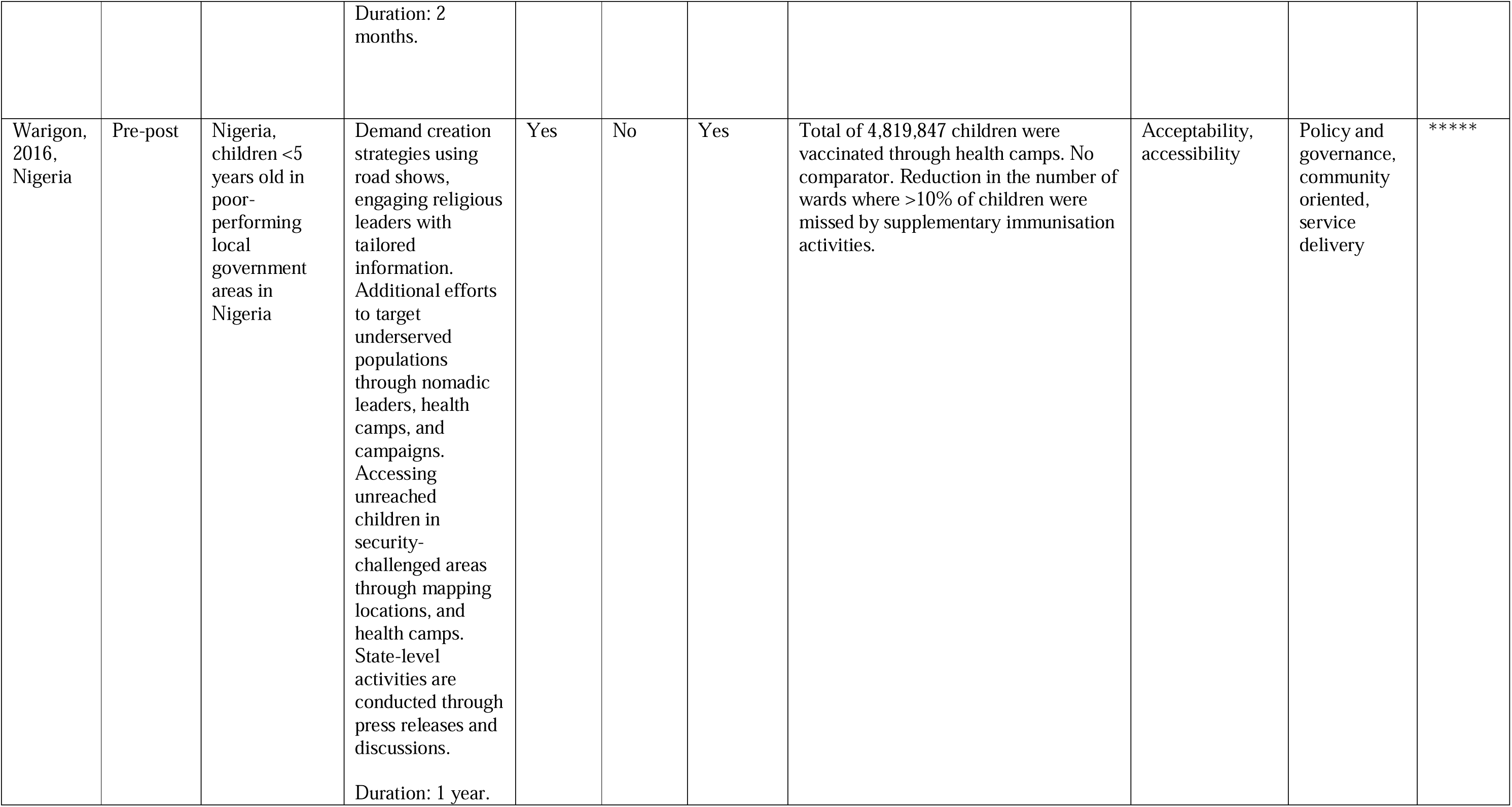
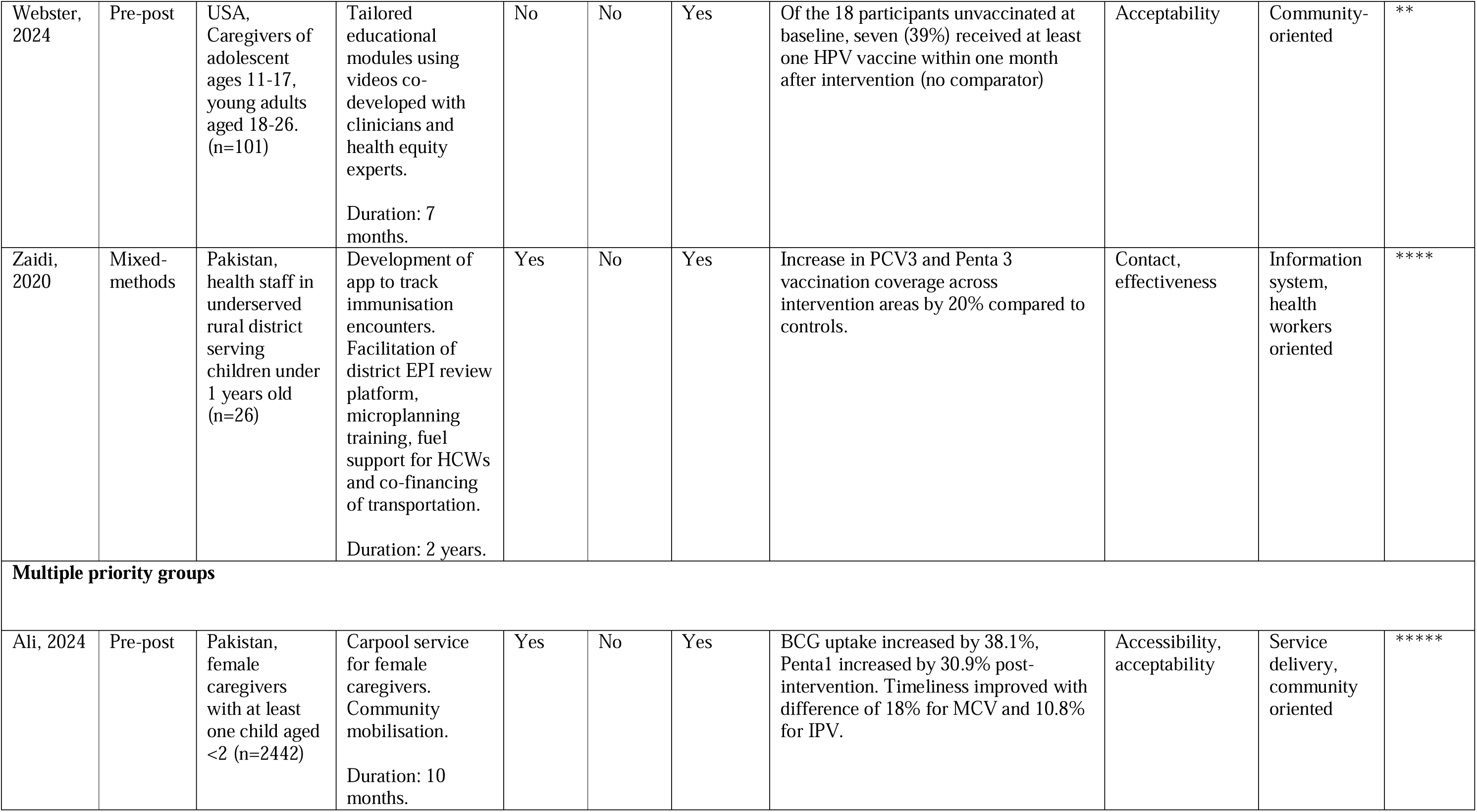
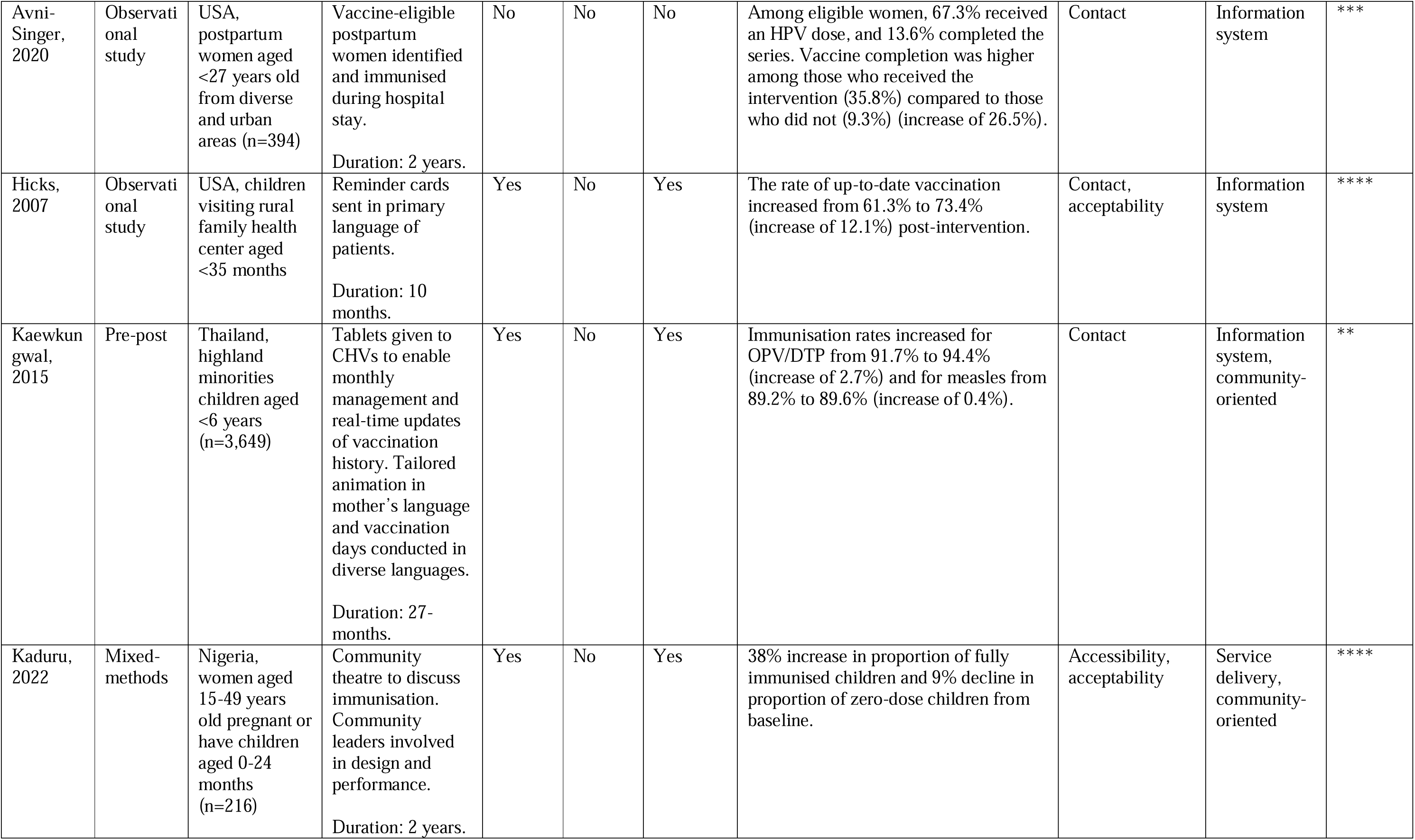
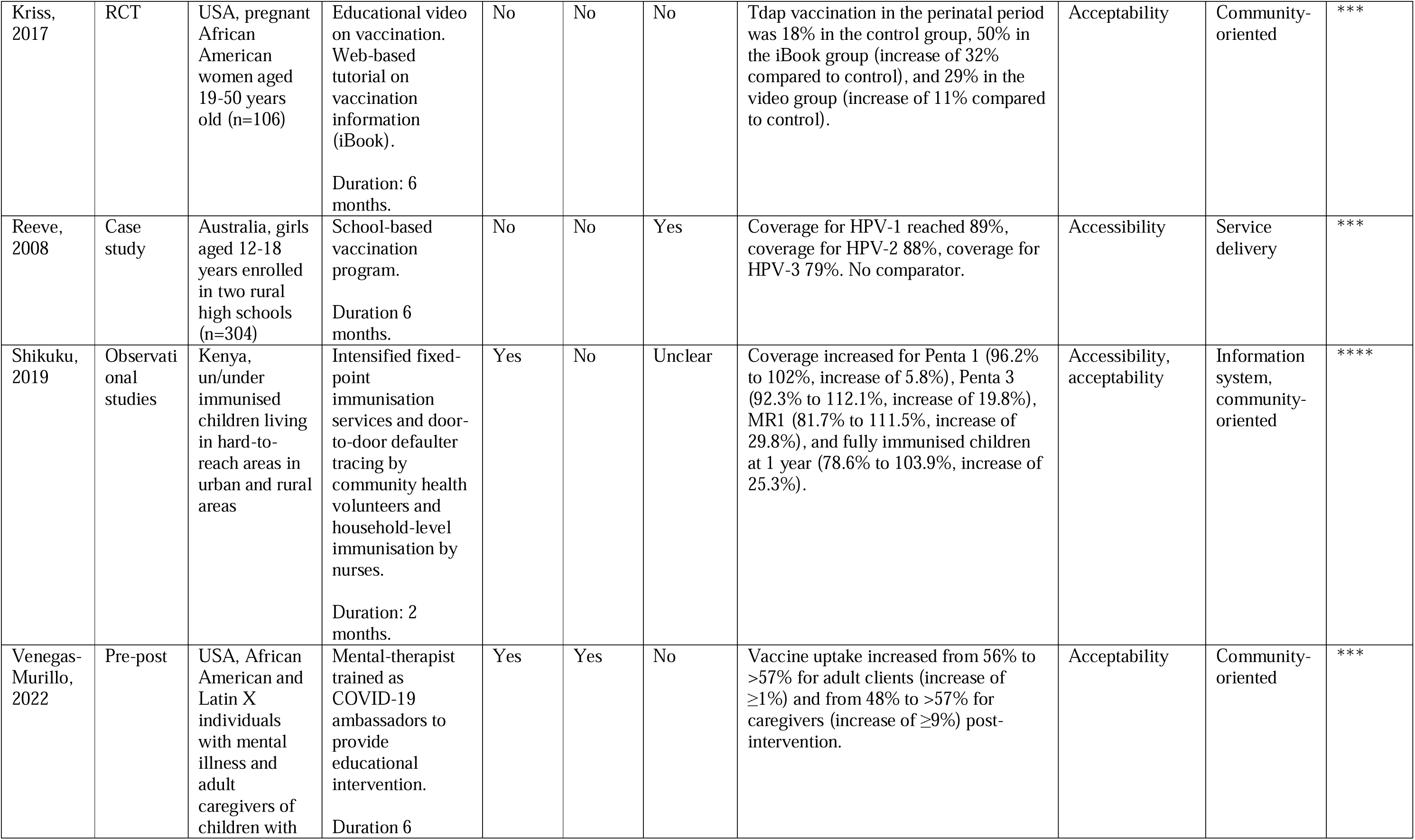
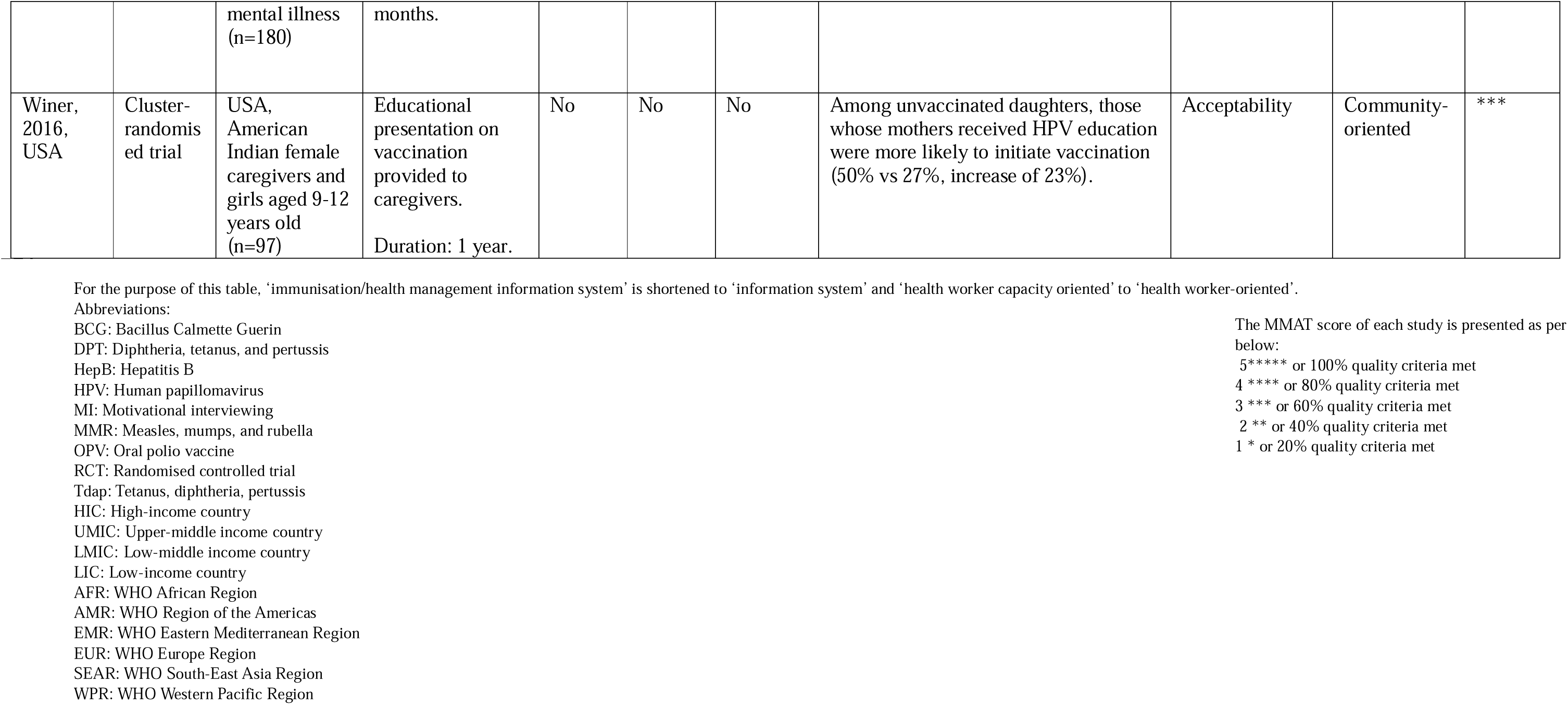
Characteristics of included studies.

| Author, year | Study design | Target population of intervention | Interventions and Duration | Multiple interventions | Outbreak | Part of routine immunisation | Key result | Tanahashi domains | Immunisation system domains | MMAT results |
| --- | --- | --- | --- | --- | --- | --- | --- | --- | --- | --- |
| **Gender** |  |  |  |  |  |  |  |  |  |  |
| Bennett, 2015 | RCT | USA, female students aged 18-26 years (n=661) | Educational website tailored to baseline survey responses.<br>Duration: 3 months | No | No | No | 8.4% of all participants initiated the vaccine series, with 42.9% of those also receiving a second dose. The proportion of participants with high knowledge about HPV vaccination increased from 32% to 50% (increase of 18%). | Acceptability | Community-oriented | ** |
| Bishop, 2021, | Observational study | South Africa, pregnant women (n=62,979) | Additional doses of vaccine added to stock and administered as part of routine care. Daily education talks given by HCWs.<br>Duration: 3 years. | Yes | No | No | Vaccine coverage at the targeted clinic reached 78.7%, with the majority of vaccinated pregnant women (93.5%) receiving vaccination on their first visit. No comparator. | Availability, acceptability | Service delivery, community-oriented | **** |
| Bucchiott y, 2021 | Observational study | France, women who gave birth (n=147 in 2011 and n=364 in 2015) | Oral and written information about pertussis is given to recipients by healthcare workers. Those unimmunised received a written prescriptions for the vaccine at discharge.<br>Duration: 5 years. | No | No | No | Postpartum vaccination rates increased from 17% to 42% (increase of 25%), while the percentage of women immunised against pertussis at the beginning of pregnancy increased from 32% to 52% (increase of 20%). | Acceptability | Community-oriented | **** |
| Buller, 2021 | RCT | USA, mothers of daughters aged 14-17 from 34 states (n=869) | Social media campaign promoting vaccination.<br>Duration: 1 year. | No | No | Yes | Uptake of the first dose of the HPV vaccine increased from 63.4% to 71.3% at 12 months (increase of 7.9%) and to 73.3% at 18 months (increase of 9.9%). Completion of the HPV vaccination increased from 50.2% to 62.5% at 12 months (increase of 12.3%) and to 65.9% at 18 months (increase of 15.7%). | Acceptability | Community-oriented | ***** |
| Cremer, 2024 | Pre-post | Switzerland, women >18 years old hospitalised for postpartum care within 28 days after delivery (n=262 in Phase 1 and n=233 in Phase 2) | Healthcare provider training through educational sessions for up-to-date vaccination information and communication techniques.<br>Duration unclear | No | No | Yes | Pertussis vaccination at baseline was 52.1% and increased to 72.7% post-intervention (20.6% increase). Influenza vaccination at baseline was 28.1% and increased to 55.6% post-intervention (27.5% increase). | Effectiveness | Health worker capacity | **** |
| Deshmukh, 2018 | Pre-post | USA, Intervention aimed at pregnant women and healthcare worker, with vaccine-eligible visits records studied (n=6,463) | Designating physician and nurse champions. Empowering nurses to recommend immunisation. Vaccination provided at no-cost and eliminating requirement for pre-vaccination pregnancy test.<br>Duration: 18 months. | Yes | No | No | The prevalence of women who completed the vaccination series was 20.3% higher, and those who initiated the series was 29.7% higher in the last month compared to before the intervention. | Acceptability, contact, accessibility | Community-oriented, information system, policy and governance | **** |
| Fontenot, 2020 | Pre-post | USA, men who have sex with men aged 18-26 years (n=42) | Development of tool co-designed with recipient. Tool consist of vaccine education, disease prevention information, health insurance enrolment, and vaccine cost assistance.<br>Duration: 7 months. | Yes | No | No | Of participants who are unvaccinated, not up to date, or did not report vaccine status, 23% utilised mHealth tool to obtain HPV vaccination. No comparator. | Acceptability | Community-oriented | *** |
| Gerend, 2021 | RCT | USA, unvaccinated sexual minority men aged 18-25 years from Chicago (n=150) | Education via text-messages on vaccination and other health practices.<br>Duration: 1 year. | Yes | No | No | HPV vaccine series initiation was higher among intervention participants (19.4%) compared to control participants (6.6%, increase of 12.8%). | Acceptability, contact | Community-oriented, service delivery | ***** |
| Kury, 2013 | Pre-post | Brazil, Parents of students eligible for HPV vaccines (n=300) | Mobile vaccination in local schools and permanent vaccination states.<br>Duration: 2 years. | No | No | Yes | 1st vaccine dose reached 53%, 90.1% and 87.9% of coverage, respectively, in 2010, 2011 and 2012. No comparator. | Accessibility | Service delivery | **** |
| Park, 2019 | Observational study | USA, healthcare workers working in hospitals with postpartum patients. Study evaluated women >26 years across two hospitals (n=387) | Nurse-based protocol given. Electronic medical record prompt reminders for nurses for each patient.<br>Duration: 6 months. | No | No | No | With nurse-based protocol, 32% received the HPV vaccine and 21% missed the opportunity. With EMR, 66% received the HPV vaccine and 20% missed the opportunity. | Contact | Information system | **** |
| Wang, 2021 | Pre-post | Hong Kong, men who have sex with men aged 18-54 years who have never received an HPV vaccination (n=350) | Online video promoting HPV vaccination for target group. Discount coupon after taking up complete doses. Text-based reminders.<br>Duration: 21 months. | Yes | No | No | 16.8% had taken up at least one dose of HPV vaccination 12 months post-intervention. | Acceptability, accessibility, contact | Community-oriented, policy and governance, Information system | **** |
| Wang, 2021, B | RCT | Hong Kong, men who have sex with men aged 18-45 years (n=624) | Web-based self-administered tutorial with peer-group discussing risk of disease and reasons to vaccinate followed by multiple-choice exercise. Motivational interviewing via telephone. Duration: 30 months. | Yes | No | No | HPV vaccination completion at 24 months was higher in the intervention group (17.3%) compared to the control group (7.2%, increase of 10.1%). | Acceptability | Community-oriented | *** |
| **Migrants and mobile population** |  |  |  |  |  |  |  |  |  |  |
| Ahmed, 2021 | Case study | Kenya, nomadic population living in two sub-counties in | Mapping and tracking of the nomadic population through lists from | Yes | No | Yes | 2,000 additional children who had not received any vaccine before from nomadic and non-nomadic areas were reached with new initiatives and vaccinated (no comparator). | Contact, acceptability, availability | Information system, community-oriented, health | *** |
|  |  | 78 settlements or nomadic hamlets (n=33,453) | local clan leaders and government administrators. Focal persons are used to mobilise residents, and community health volunteers are used as vaccinators. Tracking tools are used to monitor movement, and vaccination personnel are trained.<br>Duration: 1 year. |  |  |  |  |  | workers oriented, service delivery |  |
| Crawshaw, 2024 | Mixed-methods | UK, adolescents and adults aged >16 yo who are born outside of Western Europe, North America, Australia or New-Zealand (n=57) | Patients with uncertain/incomplete immunisation status referred to practice nurses. Nurses trained to assess the vaccination needs of migrants and administer vaccines per schedule.<br>Duration unclear. | Yes | No | Yes | 93% of participants referred to practice nurses for catch-up vaccination. 75% received at least one dose of any catch-up vaccine. Completion rates were lower, 64% for MMR and 12% completed the Td/IPV (no comparator). | Contact, effectiveness | Information system, health worker oriented | *** |
| Gomez, 2024 | Observational studies | Colombia, adults aged ≥18 years, who are born in Venezuela with Venezuelan nationality and migrated to Colombia (n=6,221) | Introduction of temporary protection status, policy communication, and decentralisation vaccination implementation.<br>Duration: 6 months. | Yes | Yes | No | A 26.2% relative increase in vaccination among individuals with irregular migration status compared to what was expected without intervention. | Accessibility, acceptability | Policy and governance, community-oriented | **** |
| Haelterm an, 199 | Pre-post | Rwanda, refugees <6 months living in two refugee camps (n=180,000 in Kibumba and 80,000 in Katala) | Vaccination campaign through setting-up vaccination posts, communication through radio, and home visits.<br>Duration: 3 months. | No | Yes | No | In Kibumba, 121,588 additional doses of vaccine were administered, covering 76% of population. No comparator. | Accessibility | Service delivery | **** |
| Kamadje u, 2015 | Observational study | Somalia, Pastoralist communities in five regions. (n=26,293) | Integrated human and animal vaccination campaigns with social mobilisation. Interpersonal communication and involvement of local community elders.<br>Duration: 2 months. | Yes | Yes | Yes | 26,293 children received OPV, 23,099 received measles vaccine. 31% of children vaccinated were zero-dose. No comparator. | Contact, availability, acceptability | Service delivery, community-oriented | ** |
| Oladeji, 2019 | Observational study | South Sudan, caregivers of vaccines-eligible children in three Outpatient Therapeutic Program, in refugee camps | Integration of immunisation into nutrition services in outpatient centres.<br>Duration: 1 year. | No | No | Yes | The number of children immunised increased post-intervention for BCG (1366 vs. 985, increase of 381), oral polio (1653 vs. 987, increase of 666), 3rd dose pentavalent (1783 vs. 1002, increase of 781), and measles (3357 vs. 2693, increase of 664). | Contact | Service delivery | *** |

| People living with disability |  |  |  |  |  |  |  |  |  |  |
| --- | --- | --- | --- | --- | --- | --- | --- | --- | --- | --- |
| Bartels, 2024 | RCT | USA, adults with mental disabilities residing in group homes (n=415) | Standard CDC-informed COVID-19 prevention protocols plus tailored peer-led vaccination testimonials, motivational interviewing, and interactive education session.<br>Duration: 12 months. | No | Yes | No | No statistically significant difference between control group and tailored group. Among unvaccinated non-White residents, tailored interventions increased vaccination rates from 14.4% to 28.6% (14.2% increase). | Acceptability | Community oriented | ** |
| Berenbrok, 2023 | Non-randomised controlled trial | USA, individuals attending 58 grocery store chain pharmacies across two groups: intervention (n=1569) and control (n=3640) | Motivational interviewing between pharmacist and patient at prescription pick-up.<br>Duration unclear. | No | No | No | HepB vaccination increased by 3.71% in the intervention group compared to the control, and 61.5% of the MI group completed the vaccination series. | Acceptability | Community-oriented | **** |
| Bock, 2016 | Pre-post | Nepal, Patients hospitalised for streptococcus pneumoniae infection (n=81) | HCW education, new medical form to track vaccinate administration. Reasons for non-administration recorded. Eligible patients vaccinated at discharge. | Yes | No | No | Vaccination rate increased from 2.5% to 42% (increase of 39.5%) post-intervention. | Effectiveness, contact | Health workers oriented, information system | **** |
|  |  |  | Duration: 2 months. |  |  |  |  |  |  |  |
| Bond, 2011 | Group-randomised evaluation | USA, patient with end-stage renal disease attending dialysis centres (n=77) | Educational seminars for HCWs, creation of action plans to improve coverage, and monthly calls between networks and centres to monitor plan implementation and vaccination coverage.<br>Duration: 1 year. | Yes | No | No | 8.9% increased in uptake in higher in intensive-intervention centres compared to standard-intervention centres. | Acceptability, contact, effectiveness | Health workers oriented, information system, service delivery | *** |
| Broderick, 2018 | Pre-post | USA, patients with rheumatoid arthritis from an outpatient centre (n=228) | Electronic Medical Based Record to alert when vaccination is due.<br>Duration: 1 year. | No | No | No | Missed opportunities for influenza vaccination decreased from 57% to 23% (reduction of 34%) post-intervention. | Contact | Information system | ***** |
| Chadwick, 2018 | Observational studies | UK, people living with HIV attending a primary care centre (n=73) | Vaccine passport to record which vaccines are provided and when boosters are required.<br>Duration: 10 months. | No | No | No | 56% improvement in immunisation coverage compared to before intervention, but full coverage not achieved. | Contact | Information system | *** |
| Christensen, 2015 | Mixed-methods | Denmark, patients with inflammatory bowel disease | Information program to healthcare workers about | No | No | No | Complete adherence increased from 5% to 26% (increase of 21%), partial adherence from 38% to 56% (increase of 18%), and complete non-adherence | Effectiveness | Health workers oriented | **** |
|  |  | (IBD) (n=130) and healthcare workers in an IBD clinic (n=53) | screening and vaccination guidelines.<br>Duration 17 months. |  |  |  | decreased from 57% to 18% (reduction of 39%). Adherence to all individual vaccinations, except HPV, increased significantly. |  |  |  |
| Cuna, 2017 | Pre-post | USA, parents of preterm infants in a neonatal intensive care unit (n=72) | Evidence-based education, recall system using electronic medical records, and standardised protocol for informed consent.<br>Duration unclear. | Yes | No | Yes | On-time vaccination increased from 36% to 82% (increase of 46%) following the intervention. | Effectiveness, contact | Health workers oriented, information system | ** |
| Ernst, 2017 | Observational study | USA, Healthcare workers using electronic alerts for infants in neonatal intensive care units (n=261 infants) | Electronic alerts to remind providers when immunisation were due for each patient.<br>Duration: 6 years. | No | No | Yes | Fully immunised rates increased from 71% to 94% (increase of 23%) after the intervention. | Contact | Information system | **** |
| Esteban-Vasallo, 2019 | Pre-post | Spain, Patients with chronic medical conditions including rare diseases (n=60,040) | Text-based reminders sent through SMS.<br>Duration: 5 months. | No | No | No | Influenza vaccine uptake was higher among those receiving reminders (9.3% vs. 7.1% in the control group, increase of 2.2%). Participants who received SMS reminders were 30% more likely to receive the influenza vaccine. | Contact | Information system | *** |
| Murthy, 2022 | Pre-post | India, caregivers of babies hospitalised in | Phone-call counselling by tele-trainers to emphasise the role | No | Yes | Yes | Immunisation uptake increased from 65.2% to 88.2% (increase of 23%) post-intervention. | Acceptability | Community-oriented | ** |
|  |  | Sick Neonatal Care Unit's across six healthcare facilities (n=3115) | of vaccines, address fears and misconceptions, and give information about vaccination location.<br>Duration: 4 months. |  |  |  |  |  |  |  |
| Stetson, 2019 | Pre-post | USA, caregivers of infants in neonatal intensive care units in a health facility (n=1242) | Quality improvement through checklist for tracking when patients are due and notifying providers. Resources on immunisation schedule and communication methods provided via intranet.<br>Duration: 6 months. | Yes | No | Yes | The fully immunised rate at the time of discharge or transfer from the NICU increased from 56% to 93% (increase of 37%) post-intervention. | Contact, acceptability, availability | Information system, community-oriented | *** |
| **Ethnic minority** |  |  |  |  |  |  |  |  |  |  |
| Abbott, 2013 | Pre-post | Australia, caregivers of children attending the Aboriginal Medical Service Western Sydney (n=505) | Calendar with list of diseases targeted in immunisation schedule with Indigenous artwork. Short health promotion messages. Duration: 18 months with 2- | Yes | No | Yes | Average delay for vaccination in intervention group was 0.6 months after the due date, compared to 3.3 months in control group. | Acceptability | Community-oriented | ** |
|  |  |  | year post-intervention period. |  |  |  |  |  |  |  |
| Lieu, 2022 | RCT | USA, Black and Latino adults aged <65 years attending primary care clinics (n=8287) | Electronic secure message and/or mail outreach using culturally tailored content.<br>Duration: 2 months. | Yes | Yes | No | COVID-19 vaccination rates were higher in the intervention group (24.0%) compared to usual care (21.7%, increase of 2.3%). | Acceptability | Community-oriented | **** |
| Marquez, 2021 | Pre-post | USA, LatinX adults living in a community, (n=11,098) | Community mobilisation, demand generation activities. Community-based and client-centred vaccination site. Social networks leveraged to increase uptake.<br>Duration: 4 months. | Yes | No | No | 91.7% received at least one dose on site. 5.7% of eligible individuals received at least one vaccine dose at neighbourhood site, including 11.9% of LatinX residents. | Accessibility, acceptability | Community-oriented, service delivery | **** |
| Tamura, 2022 | Case study | Japan, foreigners who identify as Muslims who attend a mosque | Collaboration between mosque and private clinic for vaccination site near mosque. Multilingual services are being offered. Gender-separated waiting areas. Duration: 6 months. | No | No | No | 5000 extra doses administered. Linking of unvaccinated people to clinic even after finishing vaccination at mosque. No comparator. | Acceptability | Community-oriented | ***** |
| **Urban/rural** |  |  |  |  |  |  |  |  |  |  |
| Aneni, 2013 | Observational study | Namibia, caregivers of/and orphans and vulnerable children (n=1,210) | Mobile health clinics provide vaccinations to hard-to-reach populations funded and conducted through public-private partnership.<br>Duration: 8 months. | Yes | No | Yes | Outstanding immunisations decreased from 5% to 1% for all children (reduction of 4%) and from 6.5% to 1% for the longitudinal group (reduction of 5.5%). | Accessibility, availability | Service delivery, policy and governance | *** |
| Akwataghibe, 2021 | Mixed-methods | Nigeria, community members, health workers, government officials, and caregivers (children n=213, caregivers n=210) | Participatory action research facilitating implementation of local solutions to barriers against immunisation.<br>Duration: 2 years. | No | No | Yes | Fully immunised children increased from 60.7% to 90.9% (increase of 30.2%) post-intervention. | Acceptability | Community-oriented | *** |
| Banerjee, 2010 | RCT | India, caregivers of children aged 1-3 in one of 134 rural villages (n=1640) | Monthly immunisation camps in villages. Incentive of 1kg of raw lentils per immunisation and metal plates on completion of full immunisation.<br>Duration: 32 months. | Yes | No | Yes | Full immunisation was highest with immunisation camps and incentives (144/382), compared to monthly immunisation camps only (68/379) and the control group (50/860). | Acceptability, accessibility | Service delivery, policy and governance | **** |
| Brodie, 2018, USA | Pre-post | USA, healthcare workers working in a socio-economically disadvantaged urban primary care clinic with patients of children aged 9-13 years (n=86) | Educational sessions and weekly reminders for HCWs via email on vaccine information and how to appropriately recommend vaccine.<br>Duration: 16 months. | Yes | No | Yes | Immunisation rates increased from 56% to 84% (increase of 28%) post-intervention, while missed opportunities in 11–13-year-olds decreased from 63% to 18% (reduction of 45%) post-intervention. | Effectiveness, contact | Health workers oriented, information system | * |
| Chauhadr y, 2024 | Pre-post | Paksitan, caregivers of children <5yo who have not received polio vaccination (n=122,950 at baseline and n=133,996 at endline) | Mobile health camps located within zero-dose clusters. Use of local influencers to build trust, raise awareness, and planning and promoting health camps.<br>Duration unclear. | Yes | No | Yes | Immunisation coverage increased from 39.4% at baseline to 59.6% after intervention (20.2% increase). Zero-dose prevalence decreased from 17.1% at baseline to 6.8% after intervention (10.3% decrease) | Accessibility, acceptability | Community-oriented, service delivery | *** |
| Eze, 2015 | RCT | Nigeria, adult caregivers of children <1 years old | SMS reminder with concurrent weekly data collection. Recall messages sent to defaulters.<br>Duration: 1 year. | No | No | Yes | Immunisation coverage was 8.7% higher in the intervention group than in the control, and the intervention group received the DPT3 vaccine 1.5 times earlier. | Contact | Information system | ***** |
| Kepka, 2021 | Pre-post | USA, caregivers, HCWs working in clinic and patients aged 11-26 years old due for HPV vaccination (n=402) | Healthcare team trained and educated on vaccination given. Reminder messages sent to recipient.<br>Duration: 19 months. | Yes | Yes | No | Missed opportunities for HPV vaccination declined from 21.6% to 8.1% (reduction of 13.5%) post-intervention. | Effectiveness, contact | Health workers oriented, information system | *** |
| Mupere, 2020, Uganda | Observational studies | Uganda, unreached and under-immunised children aged 0-59 months (n=191,223) | Family health days programs in places of worship as entry points to communities and points of service delivery for vaccination.<br>Duration: 16-months. | No | No | Yes | Increase coverage of 126% and 144% for measles and 103% and 122% for DTP3. | Acceptability, accessibility | Community-oriented | **** |
| Prinja, 2009, India | Observational study | India, children in rural communities aged <18 years old (n=5213) | Decentralisation of planning and management of immunisation services. Involvement of community volunteers. | Yes | No | Yes | In post-intervention cohort, 70% received third DPT dose before the age of 6 months. | Effectiveness | Policy and governance, community-oriented | **** |
|  |  |  | Duration: 18 months. |  |  |  |  |  |  |  |
| Rosenberg, 1995 | Mixed methods | USA, families with hard to reach children aged <5 years | Collaboration with community organisations for the use of community residents as outreach workers. Face-to-face communication. Assistance with using healthcare system.<br>Duration: 9 months. | Yes | Yes | Yes | The percentage of children up-to-date with OPV, DPT, and MMR increased to 73% after 9 months. | Acceptability | Community-oriented | ***** |
| Sasaki, 2011 | Observational studies | Zambia, caregivers with at least one or more children <2 years old (n=247 in 1999-2001 and 268 in 2003-2005) | Outreach immunisation services delivered at community spaces.<br>Duration 2 years. | No | No | Yes | Immunisation coverage increased for DPT3 from 75.7% to 87.3% (increase of 11.6%) and for measles from 66.8% to 76.1% (increase of 9.3%). | Accessibility | Service delivery | **** |
| Shah, 2010 | Observational study | India, unvaccinated children <5 years old in areas of mass transit (n=360,937) | Vaccination conducted at mass transit sites. Muslim vaccinators added to teams at some sites. | Yes | No | Yes | Immunisation coverage increased by 5.3% at supplementary immunisation activity sites. Transit site vaccinations increased by 18.7% in the two intervention blocks. | Accessibility, acceptability | Service delivery, health workers oriented | ** |
|  |  |  | Duration: 2 months. |  |  |  |  |  |  |  |
| Warigon, 2016, Nigeria | Pre-post | Nigeria, children <5 years old in poor-performing local government areas in Nigeria | Demand creation strategies using road shows, engaging religious leaders with tailored information. Additional efforts to target underserved populations through nomadic leaders, health camps, and campaigns. Accessing unreached children in security-challenged areas through mapping locations, and health camps. State-level activities are conducted through press releases and discussions.<br>Duration: 1 year. | Yes | No | Yes | Total of 4,819,847 children were vaccinated through health camps. No comparator. Reduction in the number of wards where >10% of children were missed by supplementary immunisation activities. | Acceptability, accessibility | Policy and governance, community oriented, service delivery | ***** |
| Webster, 2024 | Pre-post | USA, Caregivers of adolescent ages 11-17, young adults aged 18-26. (n=101) | Tailored educational modules using videos co-developed with clinicians and health equity experts.<br>Duration: 7 months. | No | No | Yes | Of the 18 participants unvaccinated at baseline, seven (39%) received at least one HPV vaccine within one month after intervention (no comparator) | Acceptability | Community-oriented | ** |
| Zaidi, 2020 | Mixed-methods | Pakistan, health staff in underserved rural district serving children under 1 years old (n=26) | Development of app to track immunisation encounters. Facilitation of district EPI review platform, microplanning training, fuel support for HCWs and co-financing of transportation.<br>Duration: 2 years. | Yes | No | Yes | Increase in PCV3 and Penta 3 vaccination coverage across intervention areas by 20% compared to controls. | Contact, effectiveness | Information system, health workers oriented | **** |
| **Multiple priority groups** |  |  |  |  |  |  |  |  |  |  |
| Ali, 2024 | Pre-post | Pakistan, female caregivers with at least one child aged <2 (n=2442) | Carpool service for female caregivers. Community mobilisation.<br>Duration: 10 months. | Yes | No | Yes | BCG uptake increased by 38.1%, Penta1 increased by 30.9% post-intervention. Timeliness improved with difference of 18% for MCV and 10.8% for IPV. | Accessibility, acceptability | Service delivery, community oriented | ***** |
| Avni-Singer, 2020 | Observational study | USA, postpartum women aged <27 years old from diverse and urban areas (n=394) | Vaccine-eligible postpartum women identified and immunised during hospital stay.<br>Duration: 2 years. | No | No | No | Among eligible women, 67.3% received an HPV dose, and 13.6% completed the series. Vaccine completion was higher among those who received the intervention (35.8%) compared to those who did not (9.3%) (increase of 26.5%). | Contact | Information system | *** |
| Hicks, 2007 | Observational study | USA, children visiting rural family health center aged <35 months | Reminder cards sent in primary language of patients.<br>Duration: 10 months. | Yes | No | Yes | The rate of up-to-date vaccination increased from 61.3% to 73.4% (increase of 12.1%) post-intervention. | Contact, acceptability | Information system | **** |
| Kaewkungwal, 2015 | Pre-post | Thailand, highland minorities children aged <6 years (n=3,649) | Tablets given to CHVs to enable monthly management and real-time updates of vaccination history. Tailored animation in mother's language and vaccination days conducted in diverse languages.<br>Duration: 27-months. | Yes | No | Yes | Immunisation rates increased for OPV/DTP from 91.7% to 94.4% (increase of 2.7%) and for measles from 89.2% to 89.6% (increase of 0.4%). | Contact | Information system, community-oriented | ** |
| Kaduru, 2022 | Mixed-methods | Nigeria, women aged 15-49 years old pregnant or have children aged 0-24 months (n=216) | Community theatre to discuss immunisation. Community leaders involved in design and performance.<br>Duration: 2 years. | Yes | No | Yes | 38% increase in proportion of fully immunised children and 9% decline in proportion of zero-dose children from baseline. | Accessibility, acceptability | Service delivery, community-oriented | **** |
| Kriss, 2017 | RCT | USA, pregnant African American women aged 19-50 years old (n=106) | Educational video on vaccination. Web-based tutorial on vaccination information (iBook).<br>Duration: 6 months. | No | No | No | Tdap vaccination in the perinatal period was 18% in the control group, 50% in the iBook group (increase of 32% compared to control), and 29% in the video group (increase of 11% compared to control). | Acceptability | Community-oriented | *** |
| Reeve, 2008 | Case study | Australia, girls aged 12-18 years enrolled in two rural high schools (n=304) | School-based vaccination program.<br>Duration 6 months. | No | No | Yes | Coverage for HPV-1 reached 89%, coverage for HPV-2 88%, coverage for HPV-3 79%. No comparator. | Accessibility | Service delivery | *** |
| Shikuku, 2019 | Observational studies | Kenya, un/under immunised children living in hard-to-reach areas in urban and rural areas | Intensified fixed-point immunisation services and door-to-door defaulter tracing by community health volunteers and household-level immunisation by nurses.<br>Duration: 2 months. | Yes | No | Unclear | Coverage increased for Penta 1 (96.2% to 102%, increase of 5.8%), Penta 3 (92.3% to 112.1%, increase of 19.8%), MR1 (81.7% to 111.5%, increase of 29.8%), and fully immunised children at 1 year (78.6% to 103.9%, increase of 25.3%). | Accessibility, acceptability | Information system, community-oriented | **** |
| Venegas-Murillo, 2022 | Pre-post | USA, African American and Latin X individuals with mental illness and adult caregivers of children with | Mental-therapist trained as COVID-19 ambassadors to provide educational intervention.<br>Duration 6 | Yes | Yes | No | Vaccine uptake increased from 56% to >57% for adult clients (increase of $\geq 1\%$) and from 48% to >57% for caregivers (increase of $\geq 9\%$) post-intervention. | Acceptability | Community-oriented | *** |
|  |  | mental illness<br>(n=180) | months. |  |  |  |  |  |  |  |
| Winer,<br>2016,<br>USA | Cluster-<br>randomis<br>ed trial | USA,<br>American<br>Indian female<br>caregivers and<br>girls aged 9-12<br>years old<br>(n=97) | Educational<br>presentation on<br>vaccination<br>provided to<br>caregivers.<br>Duration: 1 year. | No | No | No | Among unvaccinated daughters, those<br>whose mothers received HPV education<br>were more likely to initiate vaccination<br>(50% vs 27%, increase of 23%). | Acceptability | Community-oriented | *** |

For the purpose of this table, 'immunisation/health management information system' is shortened to 'information system' and 'health worker capacity oriented' to 'health worker-oriented'.
BCG: Bacillus Calmette Guerin
DPT: Diphtheria, tetanus, and pertussis
HepB: Hepatitis B
HPV: Human papillomavirus
MI: Motivational interviewing
MMR: Measles, mumps, and rubella
OPV: Oral polio vaccine
RCT: Randomised controlled trial

Across the 59 studies, we identified 67 unique interventions classified into 25 pro-equity strategies (Supplementary Table 1) and five immunisation system domains (Supplementary Table 2) through inductive thematic analyses. Community-oriented strategies were most frequently used to address immunisation inequity (33/59; 56%) while policy and governance were least frequently used (7/59; 12%). When applying the Tanahashi framework (Supplementary Table 3), most studies described strategies addressing acceptability (37/59; 63%). In contrast, only 5 (8%) studies focused on availability and 10 (17%) on effectiveness (Table 3).

**Table 3.**
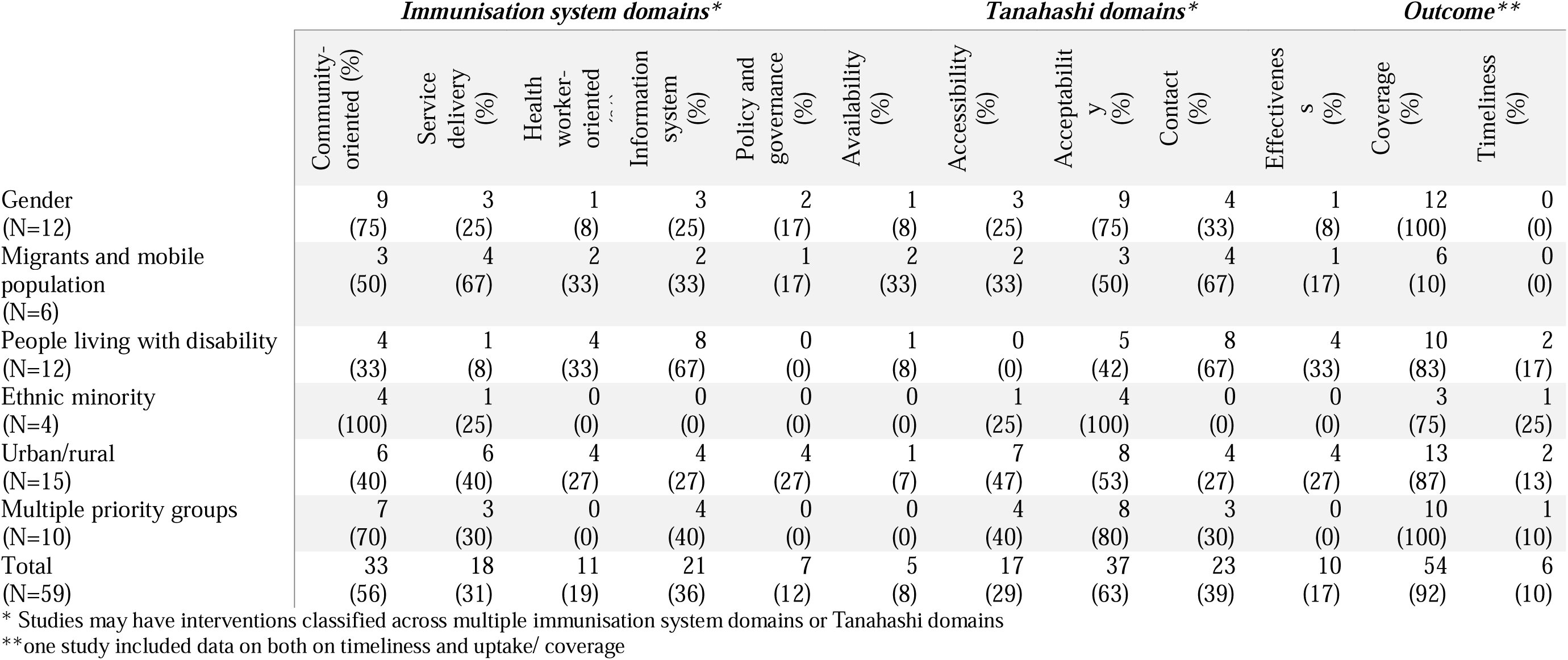
Classification of pro-equity strategies by immunisation system, Tanahashi domains, and outcome in the included studies.

|  | *Immunisation system domains\** |  |  |  |  | *Tanahashi domains\** |  |  |  | *Outcome\*\** |  |  |
| --- | --- | --- | --- | --- | --- | --- | --- | --- | --- | --- | --- | --- |
|  | Community-oriented (%) | Service delivery (%) | Health worker-oriented (%) | Information system (%) | Policy and governance (%) | Availability (%) | Accessibility (%) | Acceptability (%) | Contact (%) | Effectiveness (%) | Coverage (%) | Timeliness (%) |
| Gender (N=12) | 9 (75) | 3 (25) | 1 (8) | 3 (25) | 2 (17) | 1 (8) | 3 (25) | 9 (75) | 4 (33) | 1 (8) | 12 (100) | 0 (0) |
| Migrants and mobile population (N=6) | 3 (50) | 4 (67) | 2 (33) | 2 (33) | 1 (17) | 2 (33) | 2 (33) | 3 (50) | 4 (67) | 1 (17) | 6 (10) | 0 (0) |
| People living with disability (N=12) | 4 (33) | 1 (8) | 4 (33) | 8 (67) | 0 (0) | 1 (8) | 0 (0) | 5 (42) | 8 (67) | 4 (33) | 10 (83) | 2 (17) |
| Ethnic minority (N=4) | 4 (100) | 1 (25) | 0 (0) | 0 (0) | 0 (0) | 0 (0) | 1 (25) | 4 (100) | 0 (0) | 0 (0) | 3 (75) | 1 (25) |
| Urban/rural (N=15) | 6 (40) | 6 (40) | 4 (27) | 4 (27) | 4 (27) | 1 (7) | 7 (47) | 8 (53) | 4 (27) | 4 (27) | 13 (87) | 2 (13) |
| Multiple priority groups (N=10) | 7 (70) | 3 (30) | 0 (0) | 4 (40) | 0 (0) | 0 (0) | 4 (40) | 8 (80) | 3 (30) | 0 (0) | 10 (100) | 1 (10) |
| Total (N=59) | 33 (56) | 18 (31) | 11 (19) | 21 (36) | 7 (12) | 5 (8) | 17 (29) | 37 (63) | 23 (39) | 10 (17) | 54 (92) | 6 (10) |

\* Studies may have interventions classified across multiple immunisation system domains or Tanahashi domains
\*\*one study included data on both on timeliness and uptake/ coverage

Fifty-four (92%) studies reported increased uptake or coverage, of which 33 (61%) described community-oriented strategies, 18 (33%) service delivery, and 18 (33%) information strategies. Most study interventions targeted acceptability (35/54; 65%), contact (20/54; 37%), or accessibility (17/54; 31%) (Table 3). Six (10%) studies reported improved timeliness, of which 3 (50%) described community-oriented strategies and 3 (50%) information systems. Using the Tanahashi domain, 3 (50%) studies reported strategies classified as acceptability and contact, and 2 (33%) as effectiveness (Table 4).

**Table 4.** Distribution of immunisation system and Tanahashi domains by outcomes and priority groups.

|  | *Increased uptake/ coverage (N=54)\*, n (%)* | *Improved timeliness (N=6)\* n (%)* | *Total, N=59 n (%)* |
| --- | --- | --- | --- |
|  | **System Domain\*\*** |  |  |
| Community-oriented | 33 (61) | 3 (50) | 36 (61) |
| Service delivery | 18 (33) | 1 (17) | 19 (32) |
| Health worker-oriented | 9 (17) | 2 (33) | 11 (19) |
| Information system | 18 (33) | 3 (50) | 21 (36) |
| Policy and governance | 7 (13) | 0 (0) | 7 (12) |
|  | **Tanahashi Domain\*\*** |  |  |
| Availability | 5 (9) | 0 (0) | 5 (8) |
| Accessibility | 17 (31) | 1 (17) | 18 (31) |
| Acceptability | 35 (65) | 3 (50) | 38 (64) |
| Contact | 20 (37) | 3 (50) | 23 (39) |
| Effectiveness | 8 (15) | 2 (33) | 10 (17) |

\*one study included data on both timeliness and uptake/ coverage
\*\*Studies may have interventions classified across multiple immunisation system domains or Tanahashi domains

### Quality of studies

Thirty-one studies were high quality (4-5 score), 18 moderate (score of 3) and 10 lower quality (1-2 score) (Table 2). Lower scores were typically due to limitations such as unclear strategy descriptions or insufficient detail about study design, sampling or outcome measures, which reduced confidence in their findings.

### Priority group findings

Below, we present pro-equity strategies to reduce vaccine inequities by priority groups.

#### Women and other gender or sexual minorities

Twelve (20%) of the 59 studies addressed gender-related inequities (Table 1) (22–33). Most studies (8/12; 67%) targeted cisgender women (22–27,30,31), particularly pregnant and postpartum women (5/12; 42%) (23,24,26,27,31). Four studies (33%) focused on men who have sex with men, using tailored interventions such as mHealth platforms, peer-group discussions, and motivational interviewing (Table 2) (28,29,32,33). Zero studies targeted transgender or non-binary populations. Most (10/12; 83%) studies had participants aged 18–54 years, and two involved parents of adolescent girls eligible for HPV vaccination. No studies were conducted in LMICs or LICs. Six (50%) studies used multiple interventions, often combining community engagement with provider education (Table 1) (23,27–29,32,33).

The majority (9/12, 75%) of gender-related studies used community-oriented strategies to increase vaccination uptake or coverage (Table 3) (22–25,27–29,32,33). These interventions included tailored digital campaigns, motivational interviewing, and educational websites to address vaccine hesitancy and knowledge gaps. Another strategy that increased vaccination uptake or coverage is the use of information systems (3/12; 25%), for example, Electronic Medical Record (EMR)-based prompts for healthcare workers or SMS reminders for recipients and caregivers (27,31,32). None of the gender-related studies described an increase in timeliness. Most studies used interventions that address demand, with 9 (75%) studies focused on vaccine acceptability (22–25,27–29,32,33). Only one study each described interventions classified under Tanahashi’s effectiveness domain (26), or under availability (23) (Table 3).

#### Migrants and mobile populations

Six (10%) of the 59 studies focused on migrants and mobile populations, including nomadic pastoralists, refugees, and international migrants (Table 1) (34–39). Four (67%) studies targeted children and their caregivers (34,37–39), and two studies targeted adults 18 years and over (35,36). The four studies on routine immunisation used interventions such as co-delivery of human and animal vaccination amongst pastoralist communities, integration of nutrition and immunisation services, vaccination campaigns through mobile posts in refugee communities, and collaboration with community leaders (Table 1) (34,35,38,39). Most (4/6; 67%) applied multiple interventions using community health workers as vaccinators, decentralisation of planning, and vaccination campaigns (Table 1) (34–36,38).

All studies that targeted migrants and mobile populations increased vaccination uptake or coverage, but no studies described an increase in timeliness. Most (4/6; 67%) studies used service delivery strategies, including mobile clinics in remote areas, targeted vaccination campaigns, and integrated human-animal vaccination services designed for pastoralist communities (34,37–39). Community-oriented approaches (3/6; 50%) also increased vaccine uptake or coverage through community engagement and home visits (34,36,38). Only one study identified a policy and governance strategy by introducing a temporary protection status that allows migrants access to free vaccination in Colombia (36) When using the Tanahashi framework, 4 (67%) studies described interventions classified under contact (34,35,38,39), and 3 (50%) under acceptability (34,36,38). These interventions address demand and utilisation of immunisation services. However, only one study describes interventions classified in the effectiveness domain aimed at improving service quality (35) (Table 3).

#### People living with disability

Twelve (20%) of the 59 studies focused on people living with disabilities (Table 1) (40–51). Most studies (11/12; 92%) focused on individuals with physical disability or chronic health conditions, such as rheumatoid arthritis. Only one study specifically included people with intellectual disabilities in the USA (40) (Table 2). Most studies (9/12; 75%) targeted adults, while 3 (25%) studies focused on caregivers of children. Four (33%) studies used interventions to improve routine immunisation. Only 2 (17%) studies were conducted in LMICs or LICs. Four (33%) studies implemented multiple interventions, such as EMR prompts for clinicians to identify patients due for immunisation, service provider training, and standardised care protocols (Table 2) (42,43,47,51).

Most studies (8/12; 67%) used interventions that improved immunisation management information systems, such as EMR alerts, reminders, and recalls (42–45,47–49,51) (Table 3), including the two studies that reported increased timeliness as an outcome (2/12; 17%) (44,47). Other strategies that improved coverage or uptake are health worker-oriented strategies (4/12; 33%) (42,43,45,46) and community-oriented strategies (4/12; 33%) (40,41,50,51). Only one study (8%) reported interventions to improve service delivery, in conjunction with health-worker-oriented and information systems strategies (43). None used policy or governance approaches. Eight (67%) studies described interventions classified under Tanahashi’s contact domain (42–45,47–49,51), 5 (42%) under acceptability (40,41,43,50,51), and 4 (33%) under effectiveness (42,43,46,47). No studies described interventions classified under the accessibility domain (Table 3).

#### Ethnic minority

Four (7%) of the 59 studies focused on improving vaccination inequities among ethnic minorities (52–55) (Table 1). These included Latinx and African American adults in the USA (53,54), First Nations Australians (52), and ethnic minorities identifying as Muslim in Japan (Table 2) (55). Three (75%) studies focused on adults or caregivers, and only one study targeted children and routine immunisation (Table 1) (52). No studies were conducted in LMICs or LICs. Three (75%) studies implemented multiple interventions, combining culturally tailored outreach with accessible, community-centred service delivery (52–54).

All studies focused on ethnic minority populations used community-oriented strategies (Table 3). These included culturally tailored messaging, engagement through trusted community members, and collaboration with ethnic and religious institutions. Only one study identified service delivery type interventions (54). No studies reported interventions to improve health worker capacity, information systems, or policy and governance. Based on the Tanahashi framework, all studies reported interventions classified under the acceptability domain. Only one study had interventions classified under accessibility (54), and none under availability, contact, or effectiveness. Interventions in these studies primarily aimed to address demand rather than supply side determinants. Three (75%) studies reported an increase in uptake or coverage. One study reported improved timeliness by providing caregivers with an immunisation calendar designed with cultural artwork (Table 3) (52).

#### Urban and rural regions

Fifteen (25%) of the 59 studies focused on geographic disparities in immunisation (Table 1) (56–70). Most (9/15; 60%) studies targeted rural populations, particularly children under five and their caregivers in countries such as India, Nigeria, Pakistan, Uganda, and Zambia (56–58,60,62–64,68,70). Six (40%) studies focused on urban populations in areas like urban slums or economically disadvantaged areas (Table 2) (59,61,65–67,69). Most (14/15; 93%) studies focused on routine immunisation. Eleven (73%) studies were conducted in LMICs or LICs (56–58,60,61,63,64,66–68,70). Ten (67%) studies reported using multiple interventions within a single study, such as mobile clinic vaccination, reminders and recalls, incentives, and public-private partnership (Table 2) (56,58–60,62,64,65,67,68,70).

Of the nine studies focused on rural populations, 5 (56%) studies used interventions classified as community-oriented (57,60,63,64,68), and 4 (44%) each as service delivery (56,58,60,68) and policy and governance strategies (Table 3) (56,58,64,68). These studies described different interventions such as mobile clinics, school-based vaccination, and decentralised planning to increase vaccination uptake and coverage. In contrast, of the six urban studies, none described policy and governance interventions; rather, two studies described each of the following strategies - community-oriented (65,69), service delivery (66,67), health worker-oriented (59,67), information system (59,61). While interventions aimed at increasing demand were most reported, with 8 (53%) studies across both urban and rural settings reporting interventions to address acceptability, focus on supply and quality of services is also evident through interventions to address accessibility (7/15; 47%), contact (4/15; 27%), and effectiveness 4 (27%). Most (13/15; 87%) studies reported an increase in coverage or uptake, but 2 (13%) reported an increase in timeliness through interventions using collaboration with the community and healthcare worker education (62,65).

#### Multiple priority groups

Ten of the 59 studies (17%) examined intersections across the multiple priority groups (71–80) (Table 1). These included ethnic minority women, migrant children in remote areas, and female caregivers in rural and low-income communities (Table 2). The majority (8/10; 80%) targeted children and their caregivers (71,73–75,77–80). However, despite addressing more than one form of disadvantage, few studies analysed how these intersecting factors influenced access or outcomes. Five (50%) studies focused on improving routine immunisation outcomes (71,73–75,77). Only three studies (30%) were conducted in LMICs or LICs (71,75,78). Six (60%) studies implemented multiple interventions, often combining community engagement, service delivery, and digital tracking systems (Table 1) (71,73–75,78,79).

Seven (70%) studies focusing on multiple priority groups used community-oriented strategies such as peer-led education, community theatre, and culturally tailored outreach campaigns (Table 3) (71,74–76,78–80). Four (40%) studies described information system strategies (72–74,78), and 3 (30%) described service delivery strategies (71,75,77), including using postpartum tracking systems and reminders for vaccine series completion. No studies used health worker education or policy-level approaches. When using the Tanahashi framework, 8 (80%) studies reported interventions aimed at addressing acceptability bottlenecks (71,73–76,78–80), 4 (40%) targeted accessibility (71,75,77,78), and 3 (30%) focused on contact (72–74). No studies described interventions targeting availability and effectiveness bottlenecks. All studies reported an increase in vaccination uptake or coverage.

Additionally, one study demonstrated increased vaccination uptake and timeliness by implementing service delivery and community-oriented strategies, with interventions aimed at addressing accessibility and acceptability bottlenecks (Table 3) (71).

## Discussion

Our systematic review found that despite the increasing recognition of pro-equity strategies in addressing disadvantages that people and communities face due to their gender, migration status, ethnicity, disability, and place of residence, empirical evidence on strategies to improve vaccination outcomes is limited. In this review, we identified 59 studies where use of pro-equity strategies increased vaccine uptake and coverage (54 studies) and timeliness of vaccination (six studies) amongst the five priority groups: gender (12 studies), migrants and mobile population (six studies), people with disability (12 studies), ethnic minorities (four studies), urban-rural regions (15 studies), and multiple priority groups (10 studies). Across the 59 studies, 25 pro-equity strategies were identified, addressing five programmatic areas, including targeting the community (i.e. community-oriented), service delivery, health worker capacity, information systems, and policy and governance. The 25 strategies were also mapped against the Tanahashi domains to systematically examine which aspect of the health system an intervention was targeted towards (13).

Despite the largest number of zero-dose children being in LMICs (81), most studies were conducted in the USA. Community-oriented strategies that aimed to increase awareness and demand for immunisation services were commonly reported (56%) and included interventions such as community engagement, collaboration with local communities, caregiver education and social media (22–25,27–29,32–34,36,38,40,41,50–55,57,60,63–65,68,69,71,73–76,78–80). Using the Tanahashi framework, 63% of studies addressed acceptability, which aimed to increase the number of people who were willing to be immunised (22–25,27–29,32–34,36,38,40,41,43,50–55,57,58,60,63,65,67–69,71,73–76,78–80). Both analytical approaches suggested that very few strategies targeted upstream barriers in the health system, i.e. through immunisation policy and governance; or availability of vaccination and immunisation services or effectiveness, which aims to improve service delivery.

As expected, we did not find a single strategy or approach universally applied. However, some approaches were more common in certain populations and settings. In addressing gender-related barriers to vaccination, education and information campaigns were frequently reported and used a diverse range of platforms (22–25,27–29,32,33). With the introduction of pregnancy vaccination in recent years, and known challenges to uptake (82), it is not unexpected that many studies (42%) focused on pregnant women. However, we found no studies that targeted vaccination in transgender and non-binary populations. Considering the increased focus on life course vaccination, use of vaccines for epidemic control of diseases like COVID-19 and MPOX, and the increased risk of these populations both from lack of health services perspective and high-risk of transmission in the case of MPOX (83), there is clearly a need to prioritise the identification of what strategies work in these populations.

Migration and population mobility are significant emerging areas in the IA2030 (1), especially due to the increasing impacts of climate change and conflict-related displacements. Most reported studies have been traditionally conducted in populations such as nomadic groups and refugees, but none looked at internal migration, which are likely to become a significant issue due to climate change.

Most (83%) studies addressing vaccination gaps in people with disability were conducted in HIC settings and used a combination of information systems, such as reminders and recalls. For example, EMR to alert when vaccination is due was used in a study of rheumatoid arthritis patients attending an outpatient clinic in the USA (44). There were limited data on attempts to address service delivery or policy/governance. Only two of the studies were conducted in LMICs, which highlights a significant gap considering the immunisation gap between children with disability compared with those without any disabilities is significant (8,84). A recent scoping review examining challenges and strategies to reduce the number of zero-dose children in children with disabilities reiterated the need for tailored interventions, especially targeting access (85). However, the study did not examine outcomes or look at a life-course approach. Studies from Fiji have found rates of vaccination to be lowest in adolescents compared with zero-dose children (8).

Data on addressing vaccination gaps in ethnic minorities were limited, and, where available, were generated in HICs and mainly applied interventions that aimed to increase vaccine acceptability focussing on the populations themselves rather than addressing bottlenecks in the health system or systemic issues within the health system which might be associated with lower vaccination rates, such as racial discrimination. As previously noted, the link between ethnic and immunisation disparities in LMICs is well documented (11), and yet, we found no intervention studies to address this inequity conducted in LMICs.

When addressing gaps in vaccination in rural and urban settings, 6 (40%) studies focused on zero-dose children living in dense urban settings (86). Traditionally, it is viewed that vaccination rates in rural settings were low, especially when compared to urban settings. However, with increasing urbanisation, data suggests that most unvaccinated and under-vaccinated children live in dense urban settings across LMICs (87). A wide range of interventions were used to address inequities in this priority group, reflective of the complex range of barriers experienced by this group. It is also likely a reflection of Gavi’s recent investment in this area, which has become increasingly important over the past decade (86,88).

Interestingly, 10 studies looked at the intersectionality across our five priority groups but mostly focused on increasing community awareness and demand for vaccination through peer-led education, community theatre, and culturally tailored outreach campaigns (71,73–76,78–80). A study from the US examined the intersectional effects of race/ethnicity and disability on flu vaccine use, which found significant race-by-disability interaction in young black adults, who had higher odds of getting vaccinated compared with white adults (89). Many studies report the negative impacts of intersectionality on health equity broadly (90–92), but not many studies examine strategies that can specifically target immunisation inequities.

The purpose of this study was not to identify a single strategy that worked across all priority groups, but to identify gaps and biases in addressing the immunisation inequities. The lack of studies (gender, ethnic minorities) and limited studies (people with disability) in LMICs raises the question of where research is being conducted and why some groups remain untargeted. Considering the burden of inequities across LMICs, there is a clear need to invest in implementation research in these settings. This review builds on existing analyses of pro-equity immunisation strategies, including those by Dadari *et al*., who found that over 75% of interventions in select Gavi-supported countries focused on addressing vaccine demand, with limited attention to supply-side issues like vaccine availability and workforce (13). Our findings align with this pattern, showing a reliance on generating vaccination demand, especially using community-oriented strategies, particularly for gender, migrant, and disability groups, while strategies addressing health systems (availability and effectiveness domains) were rarely used. Other studies, such as those by Ivanova *et al*. (15) and Ducharme et al (14), have reported similar findings with limited studies addressing upstream drivers and bottlenecks in the health system. This finding is not new, as countries, public health practitioners and academics have often critiqued the focus on quick-fixes and project-based approaches to addressing deep-entrenched inequities across immunisation, but also other health programs (93). Most included studies were small-scale and often lacked disaggregated data limiting generalisability and the ability to determine whether equity gaps were truly reduced. The MMAT results indicate that most studies were of moderate quality with only a few studies assessed as high quality. However, application of the MMAT tool enabled a rigorous appraisal of included studies.

This systematic review has multiple strengths. We conducted a broad search across multiple databases, grey literature, and reference lists without language or time restrictions. We included diverse study designs, which allowed for a rich synthesis. Classification of the pro-equity strategies using an inductive thematic analysis approach, followed by a deductive analysis using the Tanahashi framework, enabled a structured and nuanced examination of all the diverse interventions reported across the 59 studies. We were able to link strategies to specific aspects of health systems, service delivery and equity gaps. This dual approach provided a structure for meaningful comparisons across studies and priority groups. This allowed identification of strategies that worked in a nuanced manner. There were some limitations. The heterogeneity of study designs, populations, and outcome measures precluded a meta-analysis and made it challenging to assess comparative effectiveness. Similarly, excluding studies that did not report outcome data does not mean that priority groups were not targeted, but that very few studies examined the real-world effectiveness of these strategies. With the recent global health funding cuts (94,95) and diminishing health budgets, it is important to regularly evaluate and implement interventions that will reduce inequities and will be sustainable. Implementation research using a systems approach that is embedded into regular monitoring and evaluation frameworks might be one such way to address this gap (96,97).

In conclusion, we provide a comprehensive synthesis of pro-equity strategies that address vaccination gaps in population groups who are often underserved or excluded from routine immunisation programs. A range of approaches is required to address systemic barriers in the health system, including at the community level, service delivery bottlenecks, information systems, and health system policies and governance, each of which needs to be tailored and adapted for the context. Addressing persistent gaps is essential to achieve IA2030 goals and ensure that no one is left behind.

## ICMJE

All authors attest they meet the ICMJE criteria for authorship.

## CRediT authorship contribution statement

Adeline Tinessia: Investigation, Formal analysis, Writing - original draft, Writing - Review & Editing. Majdi M. Sabahelzain: Investigation, Formal analysis, Writing - Review & Editing. Catherine King: Methodology, Writing - Review & Editing. Saman Khalatbari-Soltani: Investigation, Writing - Review & Editing. Praveena Gunaratnam: Investigation Writing - Review & Editing. Ibrahim Dadari: Investigation, Writing - Review & Editing, John Carlo Lorenzo: Investigation. Soumyadeep Bhaumik: Methodology, Investigation, Writing - Review & Editing. Meru Sheel: Conceptualisation, Methodology, Investigation, Writing - original draft, Writing - Review & Editing, Supervision

## Data Availability

All data produced in the present study are available upon reasonable request to the authors

## Appendix A: PRISMA Checklist 2020

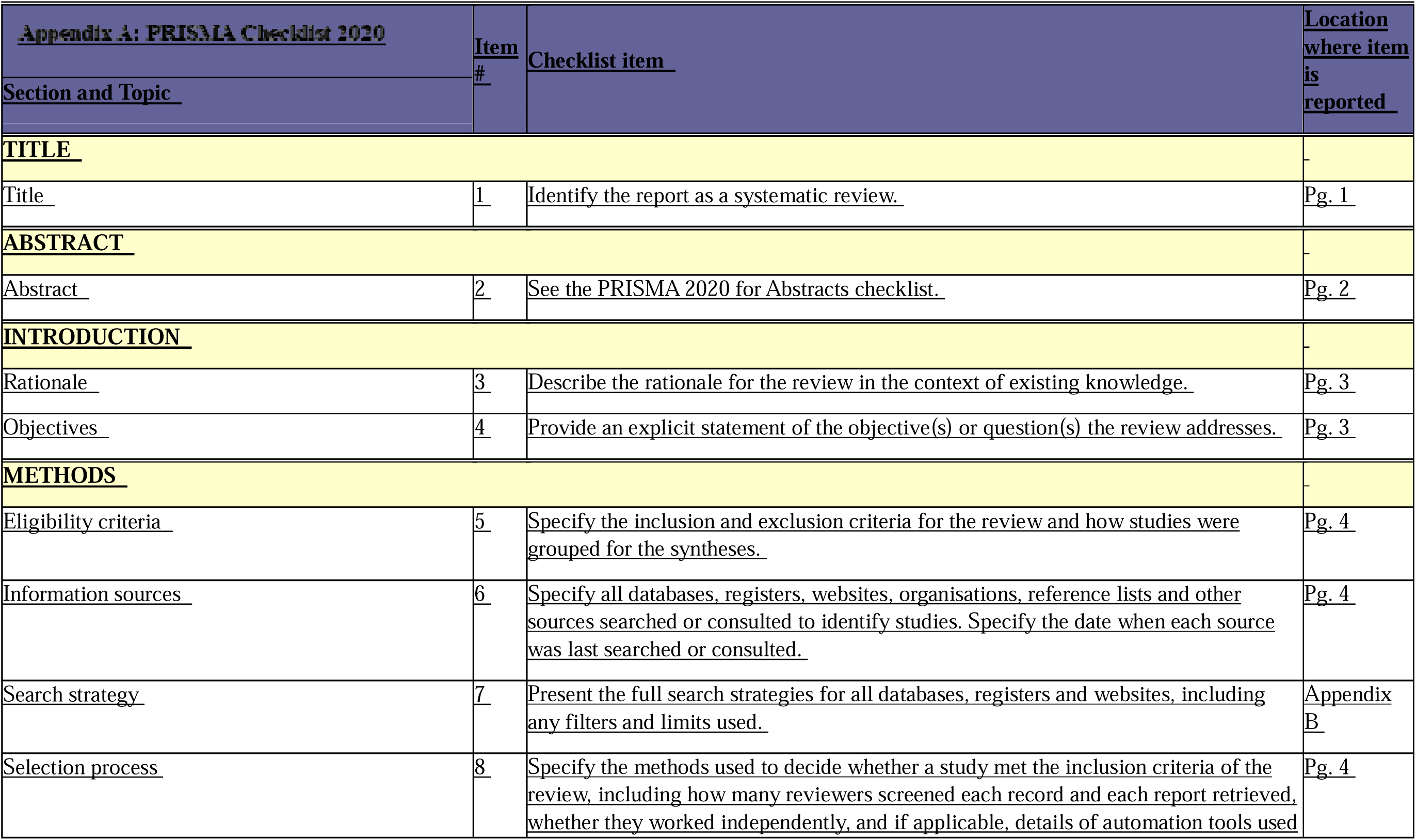
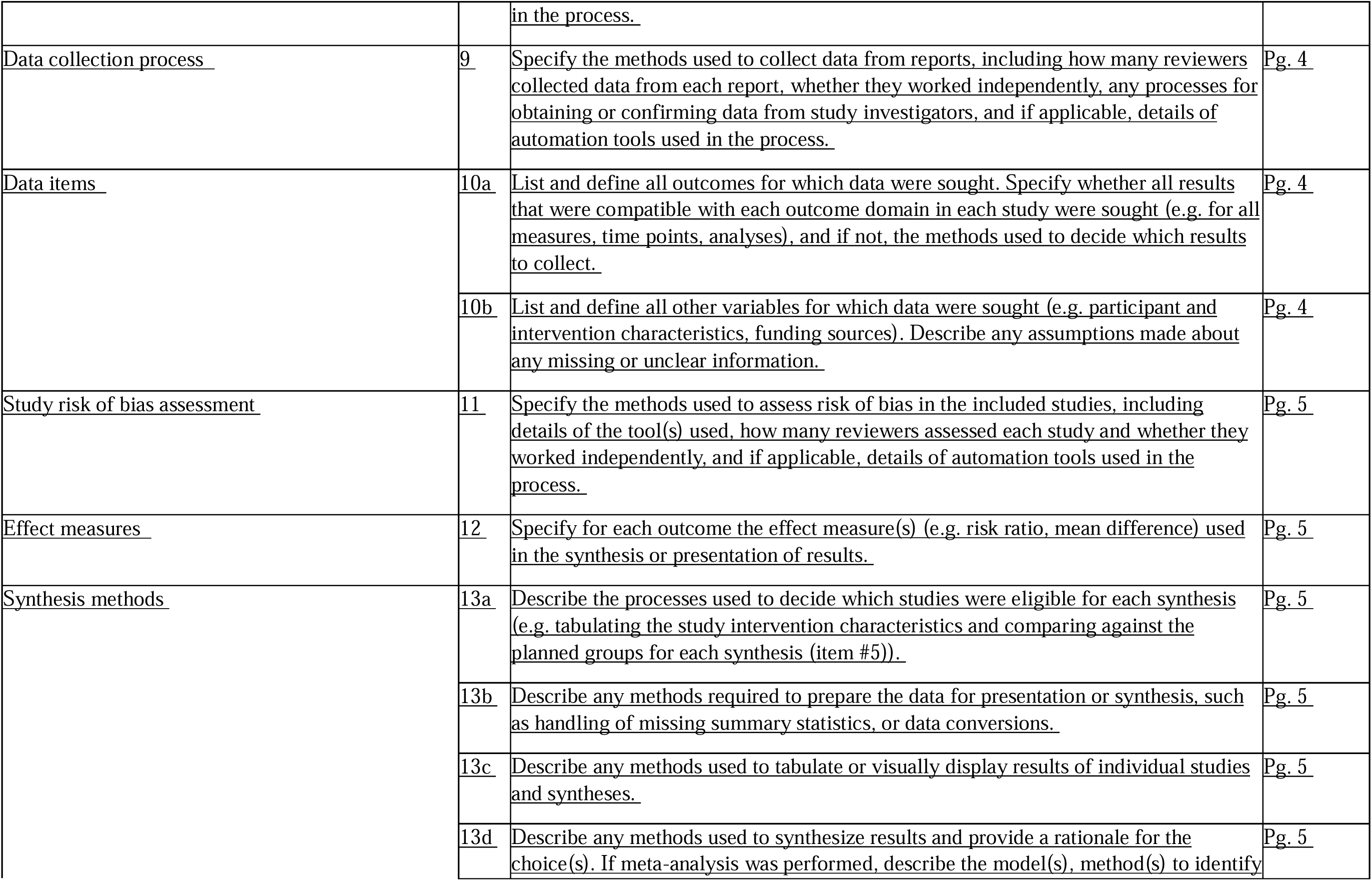
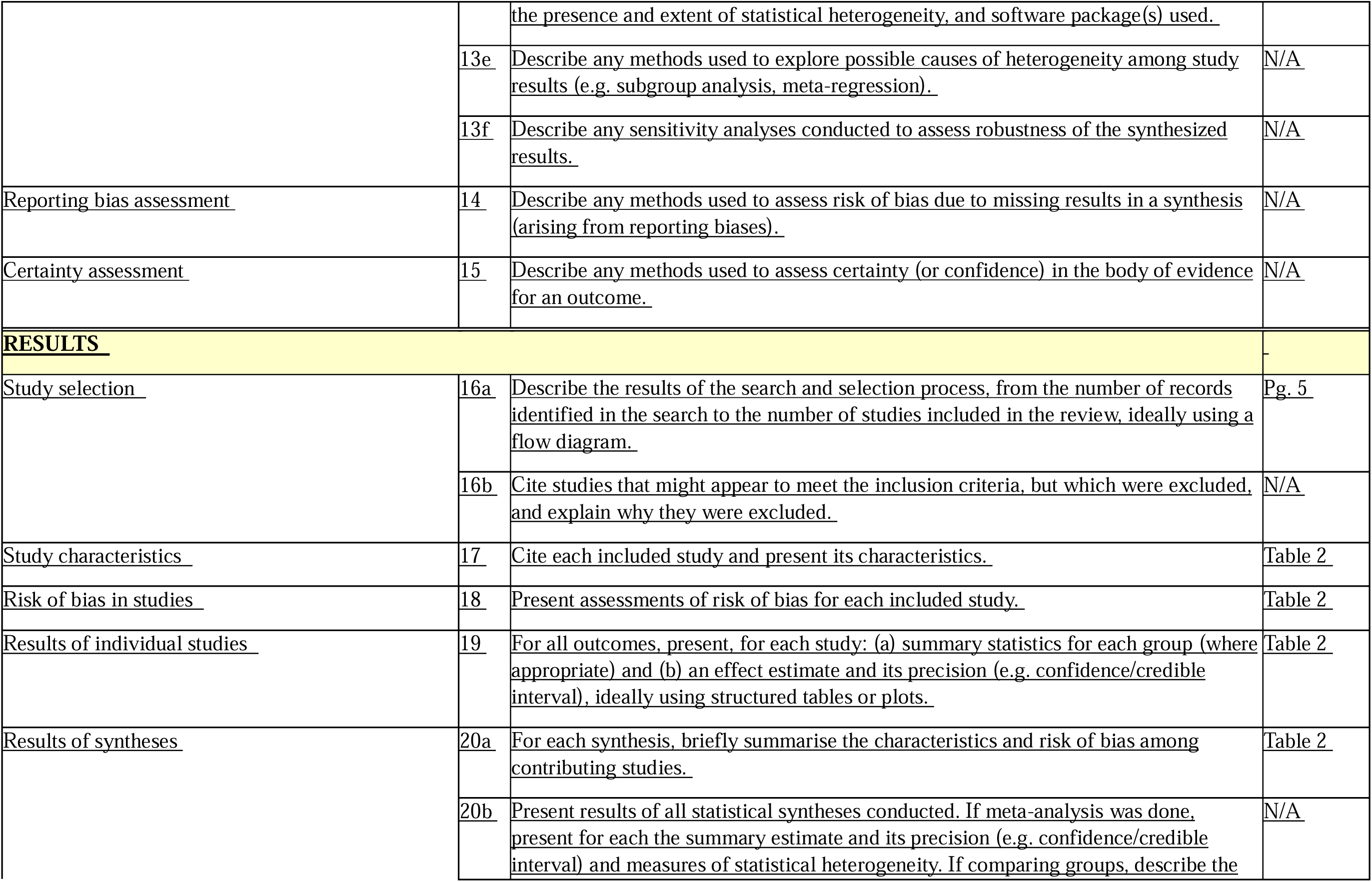
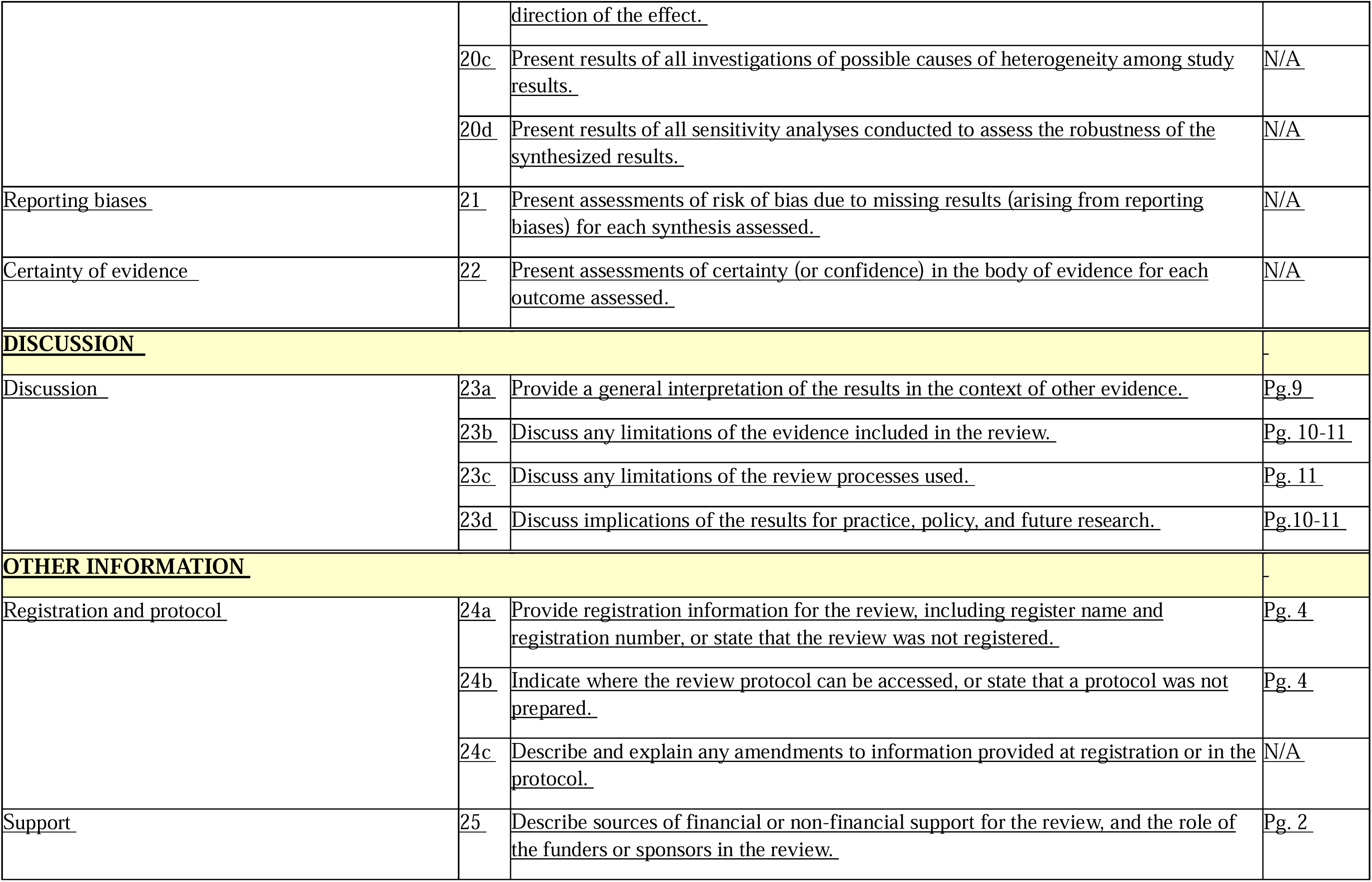
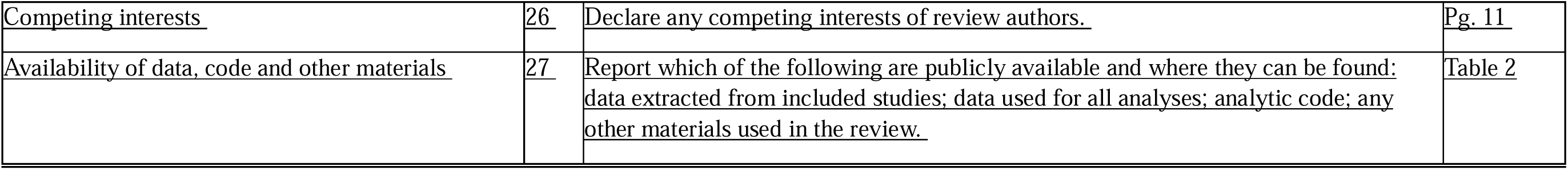

| <u>Appendix A: PRISMA Checklist 2020</u> | <u>Item #</u> | <u>Checklist item</u> | <u>Location where item is reported</u> |
| --- | --- | --- | --- |
| <u>Section and Topic</u> |  |  |  |
| **<u>TITLE</u>** |  |  |  |
| <u>Title</u> | 1 | Identify the report as a systematic review. | Pg. 1 |
| **<u>ABSTRACT</u>** |  |  |  |
| <u>Abstract</u> | 2 | See the PRISMA 2020 for Abstracts checklist. | Pg. 2 |
| **<u>INTRODUCTION</u>** |  |  |  |
| <u>Rationale</u> | 3 | Describe the rationale for the review in the context of existing knowledge. | Pg. 3 |
| <u>Objectives</u> | 4 | Provide an explicit statement of the objective(s) or question(s) the review addresses. | Pg. 3 |
| **<u>METHODS</u>** |  |  |  |
| <u>Eligibility criteria</u> | 5 | Specify the inclusion and exclusion criteria for the review and how studies were grouped for the syntheses. | Pg. 4 |
| <u>Information sources</u> | 6 | Specify all databases, registers, websites, organisations, reference lists and other sources searched or consulted to identify studies. Specify the date when each source was last searched or consulted. | Pg. 4 |
| <u>Search strategy</u> | 7 | Present the full search strategies for all databases, registers and websites, including any filters and limits used. | Appendix B |
| <u>Selection process</u> | 8 | Specify the methods used to decide whether a study met the inclusion criteria of the review, including how many reviewers screened each record and each report retrieved, whether they worked independently, and if applicable, details of automation tools used <u>in the process.</u> | Pg. 4 |
| <u>Data collection process</u> | 9 | <u>Specify the methods used to collect data from reports, including how many reviewers collected data from each report, whether they worked independently, any processes for obtaining or confirming data from study investigators, and if applicable, details of automation tools used in the process.</u> | <u>Pg. 4</u> |
| <u>Data items</u> | 10a | <u>List and define all outcomes for which data were sought. Specify whether all results that were compatible with each outcome domain in each study were sought (e.g. for all measures, time points, analyses), and if not, the methods used to decide which results to collect.</u> | <u>Pg. 4</u> |
|  | 10b | <u>List and define all other variables for which data were sought (e.g. participant and intervention characteristics, funding sources). Describe any assumptions made about any missing or unclear information.</u> | <u>Pg. 4</u> |
| <u>Study risk of bias assessment</u> | 11 | <u>Specify the methods used to assess risk of bias in the included studies, including details of the tool(s) used, how many reviewers assessed each study and whether they worked independently, and if applicable, details of automation tools used in the process.</u> | <u>Pg. 5</u> |
| <u>Effect measures</u> | 12 | <u>Specify for each outcome the effect measure(s) (e.g. risk ratio, mean difference) used in the synthesis or presentation of results.</u> | <u>Pg. 5</u> |
| <u>Synthesis methods</u> | 13a | <u>Describe the processes used to decide which studies were eligible for each synthesis (e.g. tabulating the study intervention characteristics and comparing against the planned groups for each synthesis (item #5)).</u> | <u>Pg. 5</u> |
|  | 13b | <u>Describe any methods required to prepare the data for presentation or synthesis, such as handling of missing summary statistics, or data conversions.</u> | <u>Pg. 5</u> |
|  | 13c | <u>Describe any methods used to tabulate or visually display results of individual studies and syntheses.</u> | <u>Pg. 5</u> |
|  | 13d | <u>Describe any methods used to synthesize results and provide a rationale for the choice(s). If meta-analysis was performed, describe the model(s), method(s) to identify</u> <u>the presence and extent of statistical heterogeneity, and software package(s) used.</u> | <u>Pg. 5</u> |
|  | 13e | <u>Describe any methods used to explore possible causes of heterogeneity among study results (e.g. subgroup analysis, meta-regression).</u> | <u>N/A</u> |
|  | 13f | <u>Describe any sensitivity analyses conducted to assess robustness of the synthesized results.</u> | <u>N/A</u> |
| <u>Reporting bias assessment</u> | 14 | <u>Describe any methods used to assess risk of bias due to missing results in a synthesis (arising from reporting biases).</u> | <u>N/A</u> |
| <u>Certainty assessment</u> | 15 | <u>Describe any methods used to assess certainty (or confidence) in the body of evidence for an outcome.</u> | <u>N/A</u> |
| **<u>RESULTS</u>** |  |  | - |
| <u>Study selection</u> | 16a | <u>Describe the results of the search and selection process, from the number of records identified in the search to the number of studies included in the review, ideally using a flow diagram.</u> | <u>Pg. 5</u> |
|  | 16b | <u>Cite studies that might appear to meet the inclusion criteria, but which were excluded, and explain why they were excluded.</u> | <u>N/A</u> |
| <u>Study characteristics</u> | 17 | <u>Cite each included study and present its characteristics.</u> | <u>Table 2</u> |
| <u>Risk of bias in studies</u> | 18 | <u>Present assessments of risk of bias for each included study.</u> | <u>Table 2</u> |
| <u>Results of individual studies</u> | 19 | <u>For all outcomes, present, for each study: (a) summary statistics for each group (where appropriate) and (b) an effect estimate and its precision (e.g. confidence/credible interval), ideally using structured tables or plots.</u> | <u>Table 2</u> |
| <u>Results of syntheses</u> | 20a | <u>For each synthesis, briefly summarise the characteristics and risk of bias among contributing studies.</u> | <u>Table 2</u> |
|  | 20b | <u>Present results of all statistical syntheses conducted. If meta-analysis was done, present for each the summary estimate and its precision (e.g. confidence/credible interval) and measures of statistical heterogeneity. If comparing groups, describe the</u> <u>direction of the effect.</u> | <u>N/A</u> |
|  | 20c | <u>Present results of all investigations of possible causes of heterogeneity among study results.</u> | <u>N/A</u> |
|  | 20d | <u>Present results of all sensitivity analyses conducted to assess the robustness of the synthesized results.</u> | <u>N/A</u> |
| <u>Reporting biases</u> | 21 | <u>Present assessments of risk of bias due to missing results (arising from reporting biases) for each synthesis assessed.</u> | <u>N/A</u> |
| <u>Certainty of evidence</u> | 22 | <u>Present assessments of certainty (or confidence) in the body of evidence for each outcome assessed.</u> | <u>N/A</u> |
| **<u>DISCUSSION</u>** |  |  | - |
| <u>Discussion</u> | 23a | <u>Provide a general interpretation of the results in the context of other evidence.</u> | <u>Pg.9</u> |
|  | 23b | <u>Discuss any limitations of the evidence included in the review.</u> | <u>Pg. 10-11</u> |
|  | 23c | <u>Discuss any limitations of the review processes used.</u> | <u>Pg. 11</u> |
|  | 23d | <u>Discuss implications of the results for practice, policy, and future research.</u> | <u>Pg.10-11</u> |
| **<u>OTHER INFORMATION</u>** |  |  | - |
| <u>Registration and protocol</u> | 24a | <u>Provide registration information for the review, including register name and registration number, or state that the review was not registered.</u> | <u>Pg. 4</u> |
|  | 24b | <u>Indicate where the review protocol can be accessed, or state that a protocol was not prepared.</u> | <u>Pg. 4</u> |
|  | 24c | <u>Describe and explain any amendments to information provided at registration or in the protocol.</u> | <u>N/A</u> |
| <u>Support</u> | 25 | <u>Describe sources of financial or non-financial support for the review, and the role of the funders or sponsors in the review.</u> | <u>Pg. 2</u> |
| <u>Competing interests</u> | <u>26</u> | <u>Declare any competing interests of review authors.</u> | <u>Pg. 11</u> |
| <u>Availability of data, code and other materials</u> | <u>27</u> | <u>Report which of the following are publicly available and where they can be found:</u><br><u>data extracted from included studies; data used for all analyses; analytic code; any</u><br><u>other materials used in the review.</u> | <u>Table 2</u> |

## Appendix B: Database search strategies

### Ovid MEDLINE(R) ALL <1946->

1. exp Immunization/
2. exp Immunization Programs/
3. exp Vaccines/
4. (immunis$ or immuniz$ or vaccin$).tw.
5. 1 or 2 or 3 or 4
6. exp diversity, equity, inclusion/
7. exp Health Equity/
8. 8 equit$.tw.
9. 9 inequit$.tw.
10. (pro adj2 inequit$).tw.
11. (pro-equit$ or proequit$).tw.
12. 12 gender$.tw.
13. (woman or women$ or girl$ or female$).tw.
14. exp Pregnancy/
15. (pregnan$ or mother$).tw.
16. exp Child/
17. exp Infant/
18. (baby or babies or infant$ or toddler$ or child$ or pediatric$ or paediatric$).tw.
19. exp Adult/
20. 20 adult$.tw.
21. exp Disabled Persons/
22. (disable$ or disabilit$ or impair$).tw.
23. exp Urban Population/
24. 24 urban$.tw.
25. exp Rural Population/
26. 26 rural$.tw.
27. (remote$ or inaccess$).tw.
28. ((hard or difficult$) adj4 reach$).tw.
29. exp “Ethnic and Racial Minorities”/
30. (ethnic$ adj4 minorit$).tw.
31. exp “Transients and Migrants”/
32. (migrant$ or immigrant$ or emigrant$).tw.
33. exp Refugees/
34. 34 refugee$.tw.
35. (asylum$ adj2 seek$).tw.
36. exp “Health Disparate, Minority and Vulnerable Populations”/
37. (vulnerab$ or disadvantage$ or persecut$ or margin$ or underserv$ or displace$).tw.
38. exp Communication Barriers/
39. ((communic$ or language$) adj4 barrier$).tw.
40. exp Socioeconomic Factors/
41. 41 socioeconom$.tw.
42. exp Poverty/
43. (povert$ or impover$).tw.
44. 6 or 7 or 8 or 9 or 10 or 11 or 12 or 13 or 14 or 15 or 16 or 17 or 18 or 19 or 20 or 21 or 22 or 23
45. r 24 or 25 or 26 or 27 or 28 or 29 or 30 or 31 or 32 or 33 or 34 or 35 or 36 or 37 or 38 or 39 or 40 or
46. or 42 or 43
47. 5 and 44
48. (un-vaccin$ or unvaccin$ or un-immuni$ or unimmuni$).tw.
49. (under-vaccin$ or undervaccin$ or under-immuni$ or underimmuni$).tw.
50. (partial$ adj2 (vaccin$ or immuni$ or dose$)).tw.
51. (delay$ adj2 (vaccin$ or immuni$ or dose$)).tw.
52. (incomplet$ adj2 (vaccin$ or immuni$ or dose$)).tw.
53. (miss$ adj2 (vaccin$ or immuni$ or dose$)).tw.
54. ((miss$ or lost or loss) adj2 (opportun$ or chance$)).tw.
55. (zero adj1 dos$).tw.
56. (“zero dos$” or zero-dos$).tw.
57. (no adj1 dos$).tw.
58. (lapse$ or forget$ or forgot$).tw.
59. ((leav$ or left) adj2 behind).tw.
60. 46 or 47 or 48 or 49 or 50 or 51 or 52 or 53 or 54 or 55 or 56 or 57
61. 45 and 58
62. exp Policy/
63. exp Policy Making/
64. exp Program Development/
65. (policy or policies or program$).tw.
66. (strateg$ or tool$ or interven$ or solution$ or solv$ or approach$ or facilitat$).tw.
67. exp Health Plan Implementation/
68. 66 implement$.tw.
69. (promot$ or advanc$ or champion$ or advocat$).tw.
70. exp “Delivery of Health Care”/
71. (health$ adj2 system$).tw.
72. 70 governance$.tw.
73. 60 or 61 or 62 or 63 or 64 or 65 or 66 or 67 or 68 or 69 or 70
74. 59 and 71
75. exp Vaccination Coverage/
76. (coverage$ or uptake$ or status$ or tim$).tw.
77. 73 or 74
78. 72 and 75
79. exp animals/ not humans.sh.
80. 76 not 77

### Global Health <1910->

1. exp immunization/
2. exp immunization programmes/
3. exp vaccines/
4. (immunis$ or immuniz$ or vaccin$).tw.
5. 1 or 2 or 3 or 4
6. 6 equit$.tw.
7. exp health inequalities/
8. 8 inequit$.tw.
9. (pro adj2 inequit$).tw.
10. (pro-equit$ or proequit$).tw.
11. 11 gender$.tw.
12. exp females/
13. (woman or women$ or girl$ or female$).tw.
14. exp pregnancy/
15. (pregnan$ or mother$).tw.
16. exp children/
17. exp infants/
18. (baby or babies or infant$ or toddler$ or child$ or pediatric$ or paediatric$).tw.
19. exp adults/
20. 20 adult$.tw.
21. exp people with disabilities/
22. (disable$ or disabilit$ or impair$).tw.
23. exp urban population/
24. 24 urban$.tw.
25. exp rural population/
26. 26 rural$.tw.
27. (remote$ or inaccess$).tw.
28. ((hard or difficult$) adj4 reach$).tw.
29. exp ethnic groups/
30. (ethnic$ adj4 minorit$).tw.
31. exp migrants/
32. (migrant$ or immigrant$ or emigrant$).tw.
33. exp refugees/
34. 34 refugee$.tw.
35. (asylum$ adj2 seek$).tw.
36. (vulnerab$ or disadvantage$ or persecut$ or margin$ or underserv$ or displace$).tw.
37. ((communic$ or language$) adj4 barrier$).tw.
38. exp socioeconomics/
39. 39 socioeconom$.tw.
40. exp poverty/
41. (povert$ or impover$).tw.
42. 6 or 7 or 8 or 9 or 10 or 11 or 12 or 13 or 14 or 15 or 16 or 17 or 18 or 19 or 20 or 21 or 22 or 23
43. r 24 or 25 or 26 or 27 or 28 or 29 or 30 or 31 or 32 or 33 or 34 or 35 or 36 or 37 or 38 or 39 or 40 or 41
44. 5 and 42
45. (un-vaccin$ or unvaccin$ or un-immuni$ or unimmuni$).tw.
46. (under-vaccin$ or undervaccin$ or under-immuni$ or underimmuni$).tw.
47. (partial$ adj2 (vaccin$ or immuni$ or dose$)).tw.
48. (delay$ adj2 (vaccin$ or immuni$ or dose$)).tw.
49. (incomplet$ adj2 (vaccin$ or immuni$ or dose$)).tw.
50. (miss$ adj2 (vaccin$ or immuni$ or dose$)).tw.
51. ((miss$ or lost or loss) adj2 (opportun$ or chance$)).tw.
52. (zero adj1 dos$).tw.
53. (“zero dos$” or zero-dos$).tw.
54. (no adj1 dos$).tw.
55. (lapse$ or forget$ or forgot$).tw.
56. ((leav$ or left) adj2 behind).tw.
57. 44 or 45 or 46 or 47 or 48 or 49 or 50 or 51 or 52 or 53 or 54 or 55
58. 43 and 56
59. exp policy/
60. (policy or policies or program$).tw.
61. (strateg$ or tool$ or interven$ or solution$ or solv$ or approach$ or facilitat$).tw.
62. exp health programmes/
63. 62 implement$.tw.
64. (promot$ or advanc$ or champion$ or advocat$).tw.
65. (health$ adj2 system$).tw.
66. exp governance/
67. 66 governance$.tw.
68. 58 or 59 or 60 or 61 or 62 or 63 or 64 or 65 or 66
69. 57 and 67
70. (coverage$ or uptake$ or status$ or tim$).tw.
71. 68 and 69

### Embase Classic <1947 to 1973> Part 1 of 2, Embase <1974->

1. exp immunization/
2. exp vaccine/
3. (immunis$ or immuniz$ or vaccin$).tw.
4. 1 or 2 or 3
5. exp “diversity, equity and inclusion”/
6. exp health equity/
7. 7 equit$.tw.
8. 8 inequit$.tw.
9. (pro adj2 inequit$).tw.
10. (pro-equit$ or proequit$).tw.
11. exp gender/
12. 12 gender$.tw.
13. exp female/
14. (woman or women$ or girl$ or female$).tw.
15. exp pregnancy/
16. (pregnan$ or mother$).tw.
17. exp child/
18. (baby or babies or infant$ or toddler$ or child$ or pediatric$ or paediatric$).tw.
19. exp adult/
20. 20 adult$.tw.
21. exp disabled person/
22. (disable$ or disabilit$ or impair$).tw.
23. exp urban population/
24. 24 urban$.tw.
25. exp rural population/
26. 26 rural$.tw.
27. (remote$ or inaccess$).tw.
28. ((hard or difficult$) adj4 reach$).tw.
29. exp ethnic group/
30. (ethnic$ adj4 minorit$).tw.
31. exp migration/
32. (migrant$ or immigrant$ or emigrant$).tw.
33. exp refugee/
34. 34 refugee$.tw.
35. (asylum$ adj2 seek$).tw.
36. exp vulnerable population/
37. (vulnerab$ or disadvantage$ or persecut$ or margin$ or underserv$ or displace$).tw.
38. exp communication barrier/
39. ((communic$ or language$) adj4 barrier$).tw.
40. exp socioeconomics/
41. 41 socioeconom$.tw.
42. exp poverty/
43. (povert$ or impover$).tw.
44. 5 or 6 or 7 or 8 or 9 or 10 or 11 or 12 or 13 or 14 or 15 or 16 or 17 or 18 or 19 or 20 or 21 or 22
45. r 23 or 24 or 25 or 26 or 27 or 28 or 29 or 30 or 31 or 32 or 33 or 34 or 35 or 36 or 37 or 38 or 39 or
46. or 41 or 42 or 43
47. 4 and 44
48. (un-vaccin$ or unvaccin$ or un-immuni$ or unimmuni$).tw.
49. (under-vaccin$ or undervaccin$ or under-immuni$ or underimmuni$).tw.
50. (partial$ adj2 (vaccin$ or immuni$ or dose$)).tw.
51. (delay$ adj2 (vaccin$ or immuni$ or dose$)).tw.
52. (incomplet$ adj2 (vaccin$ or immuni$ or dose$)).tw.
53. (miss$ adj2 (vaccin$ or immuni$ or dose$)).tw.
54. ((miss$ or lost or loss) adj2 (opportun$ or chance$)).tw.
55. (zero adj1 dos$).tw.
56. (“zero dos$” or zero-dos$).tw.
57. (no adj1 dos$).tw.
58. (lapse$ or forget$ or forgot$).tw.
59. ((leav$ or left) adj2 behind).tw.
60. 46 or 47 or 48 or 49 or 50 or 51 or 52 or 53 or 54 or 55 or 56 or 57
61. 45 and 58
62. exp policy/
63. exp program development/
64. (policy or policies or program$).tw.
65. (strateg$ or tool$ or interven$ or solution$ or solv$ or approach$ or facilitat$).tw.
66. exp health care planning/
67. 65 implement$.tw.
68. (promot$ or advanc$ or champion$ or advocat$).tw.
69. exp health care delivery/
70. (health$ adj2 system$).tw.
71. 69 governance$.tw.
72. 60 or 61 or 62 or 63 or 64 or 65 or 66 or 67 or 68 or 69
73. 59 and 70
74. exp vaccination coverage/
75. (coverage$ or uptake$ or status$ or tim$).tw.
76. 72 or 73
77. 71 and 74
78. (animal$ not human$).sh.
79. 75 not 76

### SCOPUS

((((TITLE-ABS-KEY (un-vaccin* OR unvaccin* OR un-immuni* OR unimmuni* OR under-vaccin* OR undervaccin* OR under-immuni* OR underimmuni*)) OR (TITLE-ABS-KEY (((partial* W/2 (vaccin* OR immuni* OR dose*))))) OR (TITLE-ABS-KEY (((delay* W/2 (vaccin* OR immuni* OR dose*))))) OR (TITLE-ABS-KEY (((incomplet* OR miss*) W/2 (vaccin* OR immuni* OR dose*)))) OR (TITLE-ABS-KEY ((((miss* OR lost OR loss) W/2 (opportun* OR chance*))))) OR (TITLE-ABS-KEY ((zero W/1 dos*))) OR (TITLE-ABS-KEY ((“zero dos*” OR zero-dos*))) OR (TITLE-ABS-KEY ((no W/1 dos*))) OR (TITLE-ABS-KEY ((no W/1 dos*))) OR (TITLE-ABS-KEY ((((leav* OR left) W/2 behind))))) AND ((TITLE-ABS-KEY (immunis* OR immuniz* OR vaccin*)) AND ((TITLE-ABS-KEY (equit* OR inequit*)) OR (TITLE-ABS-KEY (pro W/2 inequit*)) OR (TITLE-ABS-KEY (pro-equit* OR proequit*)) OR (TITLE-ABS-KEY (gender*)) OR (TITLE-ABS-KEY (woman OR women* OR girl* OR female*)) OR (TITLE-ABS-KEY (pregnan* OR mother*)) OR (TITLE-ABS-KEY (baby OR babies OR infant* OR toddler* OR child* OR pediatric* OR paediatric*)) OR (TIT LE-ABS-KEY (adult*)) OR (TITLE-ABS-KEY (disable* OR disabilit* OR impair*)) OR (TITLE-ABS-KEY (urban* OR rural* OR remote* OR inaccess*)) OR (TITLE-ABS-KEY ((((hard OR difficult*) W/4 reach*)))) OR (TITLE-ABS-KEY (ethnic* W/4 minorit*)) OR (TITLE-ABS-KEY (migrant* OR immigrant* OR emigrant*)) OR (TITLE-ABS-KEY (refugee*)) OR (TITLE-ABS-KEY (asylum* W/2 seek*)) OR (TITLE-ABS-KEY (vulnerab* OR disadvantage* OR persecut* OR margin* OR underserv* OR displace*)) OR (TITLE-ABS-KEY ((((communic* OR language*) W/4 barrier*)))) OR (TITLE-ABS-KEY (socioeconom* OR povert* OR impover*))))) AND ((TITLE-ABS-KEY (policy OR policies OR program*)) OR (TITLE-ABS-KEY (strateg* OR tool* OR interven* OR solution* OR solv* OR approach* OR facilitat*)) OR (T ITLE-ABS-KEY (implement* OR promot* OR advanc* OR champion* OR advocat*)) OR (TITLE-ABS-KEY (health* W/2 system*)) OR (TITLE-ABS-KEY (governance*)))) AND (TITLE-ABS-KEY (coverage* OR uptake* OR status* OR tim*)) AND (EXCLUDE (EXACTKEYWORD, “N onhuman”) OR EXCLUDE (EXACTKEYWORD, “Animals”) OR EXCLUDE (EXACTKEYWO RD, “Animal”)) AND (EXCLUDE (SUBJAREA, “VETE”))

