## Supplemental Tables for "Systematic review of pro-equity strategies to improve vaccination among priority populations"

**Supplementary Table 1: Expanded code classification demonstrating the thematic coding process**

| **Unique Code derived from 124 raw codes (n=67)** | **Pro-Equity Strategy (n=25)** |
| --- | --- |
| Community-based influencers | Community engagement |
| Community-based influencers | Community engagement |
| Trusted community voices | Community engagement |
| Trusted community voices | Community engagement |
| Participatory engagement | Community engagement |
| Participatory engagement | Community engagement |
| Local partnerships | Collaboration with local community |
| Local partnerships | Collaboration with local community |
| Faith-based collaboration | Collaboration with local community |
| Faith-based collaboration | Collaboration with local community |
| NGO linkage | Collaboration with local community |
| NGO linkage | Collaboration with local community |
| Culturally tailored education | Vaccine education |
| Culturally tailored education | Vaccine education |
| School-based sessions | Vaccine education |
| School-based sessions | Vaccine education |
| Parent-targeted info | Vaccine education |
| Parent-targeted info | Vaccine education |
| Gender-sensitive HCW | HCW tailored to community |
| Gender-sensitive HCW | HCW tailored to community |
| Language-matched staff | HCW tailored to community |
| Language-matched staff | HCW tailored to community |
| Peer norm reinforcement | Positive social norm |
| Peer norm reinforcement | Positive social norm |
| Social endorsement | Positive social norm |
| Social endorsement | Positive social norm |
| Mass media outreach | Media campaign |
| Mass media outreach | Media campaign |
| Radio PSA | Media campaign |
| Radio PSA | Media campaign |
| Television ads | Media campaign |
| Television ads | Media campaign |
| Integrated service days | Providing immunisation with other services |
| Integrated service days | Providing immunisation with other services |
| One-stop clinics | Providing immunisation with other services |
| One-stop clinics | Providing immunisation with other services |
| Event-based vaccination | Event-based vaccinations |
| Event-based vaccination | Event-based vaccinations |
| Back-to-school drive | Event-based vaccinations |
| Back-to-school drive | Event-based vaccinations |
| Workplace vaccination | Onsite vaccinations |
| Workplace vaccination | Onsite vaccinations |
| School-based delivery | Onsite vaccinations |
| School-based delivery | Onsite vaccinations |
| Mobile outreach | Mobile clinic |
| Mobile outreach | Mobile clinic |
| Vaccination vans | Mobile clinic |
| Vaccination vans | Mobile clinic |
| National immunisation days | Campaign |
| National immunisation days | Campaign |
| Catch-up campaign | Campaign |
| Catch-up campaign | Campaign |
| Emergency stock supply | Provision of additional stock of vaccines |
| Emergency stock supply | Provision of additional stock of vaccines |
| Microplanning for stock | Provision of additional stock of vaccines |
| Microplanning for stock | Provision of additional stock of vaccines |
| QI audits | Service quality improvement |
| QI audits | Service quality improvement |
| Service protocols | Service quality improvement |
| Service protocols | Service quality improvement |
| Transport vouchers | Transportation to immunisation service |
| Transport vouchers | Transportation to immunisation service |
| Mobile pickup units | Transportation to immunisation service |
| Mobile pickup units | Transportation to immunisation service |
| CHW-led delivery | Community health workers as vaccinators |
| CHW-led delivery | Community health workers as vaccinators |
| Door-to-door services | Community health workers as vaccinators |
| Door-to-door services | Community health workers as vaccinators |
| Skill training | Training for HCWs |
| Skill training | Training for HCWs |
| Refresher courses | Training for HCWs |
| Refresher courses | Training for HCWs |
| EMR alerts | Notification/reminder for HCW in EIR system |
| EMR alerts | Notification/reminder for HCW in EIR system |
| Digital HCW prompts | Notification/reminder for HCW in EIR system |
| Digital HCW prompts | Notification/reminder for HCW in EIR system |
| SMS reminders | Reminder/recall to caregiver/recipient |
| SMS reminders | Reminder/recall to caregiver/recipient |
| Phone call follow-ups | Reminder/recall to caregiver/recipient |
| Phone call follow-ups | Reminder/recall to caregiver/recipient |
| Electronic registry | Implementing/the use of EIR |
| Electronic registry | Implementing/the use of EIR |
| Digital records | Implementing/the use of EIR |
| Digital records | Implementing/the use of EIR |
| Manual card updates | Improving immunisation record |
| Manual card updates | Improving immunisation record |
| Data quality checks | Improving immunisation record |
| Data quality checks | Improving immunisation record |
| Referral cards | Improving referral system |
| Referral cards | Improving referral system |
| Referral linkages | Improving referral system |
| Referral linkages | Improving referral system |
| Corporate partners | Public/private partnership |
| Corporate partners | Public/private partnership |
| NGO partnership | Public/private partnership |
| NGO partnership | Public/private partnership |
| Subnational planning | Decentralised planning |
| Subnational planning | Decentralised planning |
| Localised decision-making | Decentralised planning |
| Localised decision-making | Decentralised planning |
| Cash transfer | Incentives |
| Cash transfer | Incentives |
| Food voucher | Incentives |
| Food voucher | Incentives |
| Legal documentation | Temporary protection status |
| Legal documentation | Temporary protection status |
| Protection eligibility | Temporary protection status |
| Protection eligibility | Temporary protection status |

Raw codes (n=124) refer to the initial, granular codes generated during inductive thematic analysis of intervention descriptions, of which 67 were unique. These were iteratively reviewed, grouped, and synthesised to enhance conceptual clarity into 25 over-arching pro-equity strategies.

EIR: Electronic immunisation registry

HCW: Healthcare workers

NGO: Non-government organisation

PA: Public service announcement

QI: Quality improvement

**Supplementary Table 2. Immunisation System Domains: Thematic analysis of included studies**

| **Pro-equity Strategies** | **Definitions** | **Immunisation System Domains** |
| --- | --- | --- |
| Community engagement | Strategies aimed at increasing awareness and demand for immunisation services by working with and within communities | Community-oriented |
| Collaboration with local community |  |  |
| Vaccine education |  |  |
| HCW tailored to community |  |  |
| Positive social norm |  |  |
| Media campaign |  |  |
| Providing immunisation with other services | Strategies focused on actual provision of immunisation services by ensuring immunisation services are accessible and reached by all who need them | Service delivery |
| Event-based vaccinations |  |  |
| Onsite vaccinations |  |  |
| Mobile clinic |  |  |
| Campaign |  |  |
| Provision of additional stock of vaccines |  |  |
| Service quality improvement |  |  |
| Transportation to immunisation service |  |  |
| Community health workers as vaccinators | Strategies targeted at building capacity of frontline workers who deliver immunisation services, including vaccinators, data persons, and managers | Health worker capacity oriented |
| Training for HCWs |  |  |
| Notification/reminder for HCW in EIR system | Strategies that involve the systems used to collect, analyse, and use data related to immunisation with the goal of using the data to improve immunisation coverage, identify gaps, and plan effective interventions | Immunisation/health management information system |
| Reminder/recall to caregiver/recipient |  |  |
| Implementing/the use of EIR |  |  |
| Improving immunisation record |  |  |
| Improving referral system |  |  |
| Public/private partnership | Interventions focused on the development, implementation, and enforcement of policies, regulations, and guidelines to support immunisation programs | Policy and governance |
| Decentralised planning |  |  |
| Incentives |  |  |
| Temporary protection status |  |  |

Abbreviations:

HCW: Healthcare workers

EIR: Electronic immunisation registry

**Supplementary Table 3. Tanahashi Domains Classification based on the pro-equity strategies identified from the included studies**

| **Pro-equity strategies** | **Definitions** | **Tanahashi Domains** |
| --- | --- | --- |
| Providing immunisation with other services* | Improving availability of vaccination and immunisation services for potential recipients through increasing vaccine stocks and available immunisers | Availability |
| Implementing/the use of EIR* |  |  |
| Improving immunisation record* |  |  |
| Community health workers as vaccinators |  |  |
| Provision of additional stock of vaccines |  |  |
| Public/private partnership |  |  |
| Onsite vaccinations* | Improving the number of people who can use the service by reducing and eliminating geographical, transportation, financial, and other barriers | Accessibility |
| Campaign |  |  |
| Eliminating requirements |  |  |
| Mobile clinic |  |  |
| Event-based vaccinations |  |  |
| Temporary protection status |  |  |
| Transportation to immunisation services |  |  |
| Community engagement | Increasing the number of people who are willing to be immunised with interventions; targeted on cultural, social, and other societal factors | Acceptability |
| Collaboration with local community |  |  |
| Media campaign |  |  |
| Vaccine education |  |  |
| Positive social norm |  |  |
| Onsite vaccinations* |  |  |
| Incentives |  |  |
| Improving immunisation record* | Addressing other barriers that prevent individuals from attending/re-attending vaccination sessions | Contact |
| Implementing/the use of EIR* |  |  |
| Providing immunisation with other services* |  |  |
| Notification/reminder for HCW in EIR system |  |  |
| Reminder/recall to caregiver/recipient |  |  |
| Implementing/the use of EIR |  |  |
| Improving referral system |  |  |
| Training for HCWs | Improving the satisfactory care of those using the immunisation services | Effectiveness |
| Service quality improvement |  |  |
| Decentralised planning |  |  |

Abbreviations:

EIR: Electronic immunisation registry

HCW: Healthcare workers

* Indicate pro-equity strategies that sit within two Tanahashi domains
